# Shared genetic architecture between digestive infectious diseases and cardiovascular diseases: a genome-wide cross-trait analysis

**DOI:** 10.64898/2026.11.17.26368334

**Authors:** Jinpei Ni, Jianbo Pan, Yunqing Chen

**Author notes:** Corresponding authors: Yunqing Chen. Jianbo Pan.

## Abstract

**Background:** Digestive infectious diseases and cardiovascular diseases may share common genetic, inflammatory and immune-related mechanisms. However, the shared genetic architecture between these disease groups, the specific pleiotropic loci involved and the potential contribution of intestinal infectious diseases (IID) to cardiovascular risk remain incompletely understood.

**Methods:** We analysed publicly available genome-wide association study summary statistics for 36 binary traits of European ancestry, including 16 digestive infectious disease traits and 20 cardiovascular traits. Univariate linkage disequilibrium score regression was used to estimate SNP-based heritability and prioritise traits. Global and local genetic sharing were evaluated using cross-trait LDSC and SUPERGNOVA. Multi-trait analysis of GWAS, cross-trait sensitivity analyses, FM-summary fine-mapping and Bayesian colocalisation were used to identify and refine pleiotropic loci. Candidate genes and biological functions were investigated using MAGMA, tissue- and cell-type-specific enrichment analyses, transcriptome-wide association analysis and protein-wide association analysis. Bidirectional two-sample Mendelian randomisation and two-step mediation analyses using three independent gut microbiome GWAS resources were subsequently performed.

**Results:** Sixteen traits passed heritability quality control, yielding 39 trait pairs with significant global genetic correlations after Bonferroni correction. MTAG identified 41 candidate SNP records and 14 trait-pair-specific lead-SNP records representing eight unique SNPs; 39 candidate records and 12 lead records involved IID. Fine-mapping and colocalisation prioritised eight records representing four unique variants. Locus-based gene prioritisation retained 24 candidate genes across four pairs. Gene-set enrichment identified 43 significant trait-pair-GO term associations, and tissue enrichment identified seven significant associations. Transcriptome-wide analyses implicated 21 candidate genes, and proteome-wide analyses identified four associations involving three proteins. After false discovery rate (FDR) correction, forward MR supported associations of IID liability with cardiac arrhythmias, myocardial infarction, atrial fibrillation and flutter, and ischaemic heart disease; reverse MR supported only cardiac arrhythmias to IID. Five positive exploratory microbiota-IID-cardiovascular mediation patterns were described. Only the *Oxalobacter*-IID-cardiac arrhythmias indirect effect reached unadjusted *P* < 0.05.

**Conclusions:** Digestive infectious diseases, particularly IID, share substantial genetic architecture with cardiovascular diseases. Integrative genetic analyses identified recurrent pleiotropic loci, candidate genes and putative microbiome-mediated pathways linking IID with cardiovascular outcomes. These findings provide testable hypotheses for further functional validation and clinical investigation.

## Introduction

Cardiovascular diseases remain the leading cause of death and a major contributor to disability worldwide [1, 2]. In addition to established risk factors, inflammatory processes contribute to the development and progression of atherosclerotic cardiovascular disease [3]. In CANTOS, canakinumab at 150 mg every three months reduced recurrent cardiovascular events without lowering lipid levels [4]. A recent American College of Cardiology scientific statement highlights persistent inflammation as a contributor to residual cardiovascular risk [5]. In a large multicohort study, infections severe enough to require hospitalisation were associated with a substantially increased risk of major cardiovascular events shortly after hospital discharge, with a smaller excess risk persisting over the longer term [6].

Laboratory-confirmed influenza infection has also been associated with a transient increase in myocardial infarction risk [7]. These findings suggest that infection-related biological processes may influence cardiovascular risk beyond conventional risk factors.

The gastrointestinal tract represents a major interface between the host immune system, the gut microbiome and environmental exposures. The intestinal epithelial barrier and mucosal immune system play central roles in maintaining host-microbiome homeostasis [8, 9]. Digestive infectious diseases may disrupt epithelial integrity, alter mucosal immune responses and reshape microbial communities, thereby potentially influencing systemic inflammation, vascular biology, thrombosis and cardiac electrophysiology. Studies of cholera have implicated innate immune signalling and epithelial ion transport in inherited susceptibility to enteric infection [10]. However, previous studies have largely focused on individual pathogens, specific clinical syndromes or short-term epidemiological associations. Whether digestive infectious diseases and cardiovascular diseases share common inherited susceptibility factors across disease categories remains unclear.

Genome-wide association studies (GWAS) and summary-statistics-based genetic analyses provide an opportunity to investigate this question systematically. GWAS can characterise the polygenic architecture of complex traits, whereas summary-statistics-based methods such as LDSC can estimate SNP-based heritability and quantify genetic relationships between phenotypes [11, 12]. Linkage disequilibrium score regression (LDSC) enables the estimation of genome-wide genetic correlations while accounting for linkage disequilibrium and potential confounding in summary-level data [13]. Cross-trait approaches such as multi-trait analysis of GWAS (MTAG) can increase power to identify shared variant-level signals [14], whereas Bayesian colocalisation can assess whether association signals for two traits are consistent with a shared causal variant [15]. Integrating these approaches with fine-mapping, gene-based analyses and transcriptomic or proteomic annotation may therefore provide a more comprehensive framework for resolving shared disease mechanisms. An integrative cross-trait study of sleep traits and cardiometabolic diseases combined genetic correlation, pleiotropy, colocalisation and Mendelian randomisation analyses to characterise shared genetic determinants [16]. The gut microbiome represents a potential mechanistic link between intestinal infection and cardiovascular disease. The MiBioGen consortium analysed gut microbiome data from 18,340 individuals across 24 cohorts and identified host genetic factors associated with microbial taxa, while also demonstrating substantial heterogeneity in microbial composition across cohorts [17]. Recent metagenomic GWAS have replicated associations at established host loci and identified additional links between intestinal functions and microbial composition [18]. The HUNT study identified reproducible SNP-species associations and links to microbial functional modules [19]. Host variants at *ABO* and *FUT2* have also been linked to structural variation within gut microbial genomes [20]. A previous Mendelian randomisation study examined associations between gut microbial traits and several cardiovascular outcomes, including coronary artery disease, myocardial infarction, heart failure and atrial fibrillation; however, the evidence varied by outcome and significance threshold [21]. Nevertheless, the direction and consistency of these associations vary across microbial traits and datasets. Integrated metagenomic and metabolomic profiling has distinguished features shared across cardiometabolic diseases from signatures associated with ischaemic heart disease [22]. Experimental and translational studies have also implicated microbiota-dependent metabolites in cardiovascular biology. In particular, gut microbial metabolism of phosphatidylcholine and L-carnitine can generate trimethylamine N-oxide, which has been linked to atherosclerosis and cardiovascular risk [23–25]. Beyond TMAO, microbial imidazole propionate has been linked to human atherosclerosis and shown to promote disease in experimental models [26]. Genetic analyses have additionally provided evidence for causal relationships between gut microbial features, short-chain fatty acids and metabolic phenotypes [27].

To our knowledge, few studies have integrated disease-level genetic correlation, cross-trait pleiotropy, functional annotation, bidirectional Mendelian randomisation and gut microbiome mediation to investigate the relationship between intestinal infectious diseases and cardiovascular diseases. In particular, it remains unknown whether genetic liability to intestinal infectious diseases shares common determinants with multiple cardiovascular phenotypes, whether these signals are concentrated in specific loci or genes, and whether gut microbial traits may contribute to these relationships through IID.

In this study, we analysed GWAS summary statistics for 36 European-ancestry traits, including 16 digestive infectious disease traits and 20 cardiovascular traits. We combined univariate and cross-trait LDSC, local genetic covariance analysis, MTAG, fine-mapping, colocalisation, functional annotation, bidirectional Mendelian randomisation and gut microbiome mediation analysis. We aimed to: (i) estimate and prioritise trait-level SNP heritability; (ii) characterise global and local genetic sharing between digestive infectious diseases and cardiovascular diseases; (iii) identify and functionally annotate candidate pleiotropic variants and genes; (iv) assess potential directional causal relationships; and (v) evaluate putative gut microbiota-IID-cardiovascular disease mediation pathways (Fig. 1).

**Fig. 1.**
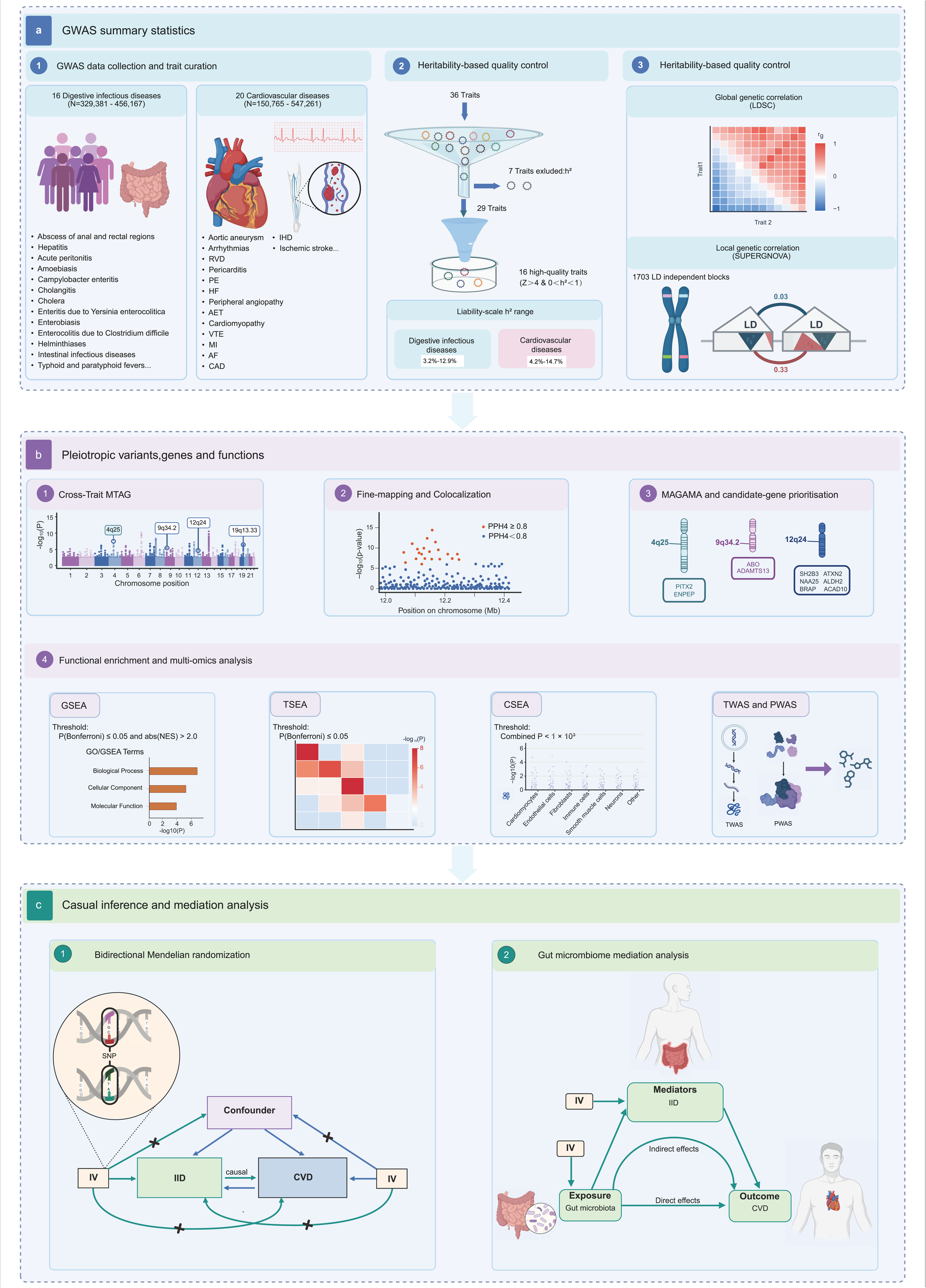
Study design and analytical workflow. Created with BioRender.com. **a** European-ancestry GWAS summary statistics for 36 traits (16 digestive infectious and 20 cardiovascular disease traits) underwent heritability-based quality control. Seven traits with negative or near-zero observed-scale heritability were excluded. Of the remaining 29, 16 high-quality traits (heritability *Z* > 4 and 0 < observed-scale h² < 1) were retained, comprising three infectious and 13 cardiovascular traits. Displayed heritability ranges are on the liability scale. Global genetic correlation and local genetic covariance were assessed using LDSC SUPERGNOVA, respectively. **b** MTAG, fine-mapping, Bayesian colocalisation and MAGMA were integrated to prioritise pleiotropic variants and genes. IID-centred functional analyses comprised GSEA, TSEA, CSEA, TWAS and PWAS. **c** Bidirectional two-sample MR evaluated potential directional IID-CVD associations. Two-step MR assessed gut microbiota-IID-CVD pathways, with IID as the mediator. Arrows show evaluated paths; crosses mark paths excluded by the MR independence and exclusion restriction assumptions. Statistical plots illustrate the workflow schematically. The CSEA combined *P* < 1 × 10 ³ threshold denotes exploratory screening. Abbreviations: AF, atrial fibrillation and flutter; AET, arterial embolism and thrombosis; CAD, coronary artery disease; CSEA, cell-type-specific enrichment analysis; CVD, cardiovascular disease; GO, Gene Ontology; GSEA, gene-set enrichment analysis; GWAS, genome-wide association study; HF, heart failure; IID, intestinal infectious diseases; IHD, ischaemic heart disease; IV, instrumental variable; LD, linkage disequilibrium; LDSC, linkage disequilibrium score regression; MAGMA, multi-marker analysis of genomic annotation; MI, myocardial infarction; MR, Mendelian randomisation; MTAG, multi-trait analysis of GWAS; NES, normalized enrichment score; PE, pulmonary embolism; PPH4, posterior probability of hypothesis H4 (shared causal variant); PWAS, proteome-wide association study; RVD, rheumatic valve diseases; SNP, single-nucleotide polymorphism; TSEA, tissue-specific enrichment analysis; TWAS, transcriptome-wide association study; VTE, venous thromboembolism; h², SNP heritability; *r*_g_, genetic correlation; *Z*, heritability *Z* score.

## Methods

### Study design and GWAS data sources

The analytical framework was informed by a previous genome-wide cross-trait study of lung and gastrointestinal diseases and adapted to investigate shared genetic architecture between digestive infectious diseases and cardiovascular diseases [28]. We used a staged genome-wide cross-trait framework to investigate shared genetic architecture between digestive infectious diseases and cardiovascular diseases and to evaluate potential directional relationships and gut microbiota-mediated pathways. Publicly available European-ancestry GWAS summary statistics were initially assembled for 36 binary traits: 16 digestive infectious disease traits and 20 cardiovascular disease traits. GWAS summary statistics were downloaded from the disease-centric Mendelian randomization database (DMRdb) [29]. Trait names, GWAS identifiers, sample sizes, case and control counts, ancestry, genome build, source studies and access information are provided in Supplementary Data 1.

Univariate linkage disequilibrium score regression (LDSC) was used to estimate SNP-based heritability and prioritise traits. Bivariate LDSC and SUPERGNOVA were then used to assess genome-wide genetic correlation and local genetic covariance, respectively (Fig. 1a). Trait pairs passing the prespecified significance and trait-quality filters were taken forward for multi-trait analysis of GWAS (MTAG). Candidate variants were evaluated using complementary cross-trait methods, fine-mapping and Bayesian colocalisation. Candidate genes and biological annotations were assessed using gene-based, enrichment, transcriptomic and proteomic analyses. Bidirectional two-sample Mendelian randomisation (MR) and two-step gut microbiome mediation analyses were subsequently performed for the IID-centred cardiovascular trait pairs (Fig. 1c).

After univariate heritability quality control, 16 traits were retained: abscess of anal and rectal regions, intestinal infectious diseases (IID; DGWAS-430) and viral hepatitis (DGWAS-1973), together with 13 cardiovascular traits listed in Supplementary Data 1. IID was the primary infectious disease phenotype for downstream (Fig. 1b).

### GWAS summary-statistic harmonisation and quality control

Genotyping, imputation and cohort-level quality control were performed by the investigators of the original GWASs. The present study used summary-level association data and performed additional data harmonisation and SNP-level preprocessing. Information on the contributing GWAS resources, sample sizes, ancestry and reported genome builds is provided in Supplementary Data 1.

Summary statistics were standardised to common fields, including SNP identifier, chromosome, base-pair position, effect and non-effect alleles, effect estimate, standard error, *P* value, effect-allele frequency where available and sample size. Odds ratios were converted to log-odds effect estimates as β = ln(OR). When an OR and its 95% confidence interval were reported without a standard error, the standard error of the log-odds estimate was calculated as [ln(upper confidence limit) − ln(lower confidence limit)]/(2 × 1.96). Binary-trait effects were analysed on the log-odds scale.

Coordinate processing preceded summary-statistic munging. The coordinate-conversion script used pyliftover with an hg38-to-hg19 mapping. Records without a returned coordinate mapping were not retained. Variants mapped to the major histocompatibility complex region on chromosome 6, from 25,000,000 to 34,000,000 bp in hg19, were excluded before the LDSC input files were prepared.

LDSC input files were prepared using the MTAG-distributed mtag_munge.py script, with explicit mappings for SNP identifiers, effect and non-effect alleles, association *P* values, beta estimates, sample sizes and allele frequencies where available. SNPs were matched to the HapMap3 reference allele file [30]. Munging included filtering of records with missing required values, invalid *P* values, duplicate SNP identifiers, non-SNP alleles, strand-ambiguous A/T or C/G allele pairs, and allele combinations incompatible with the reference file. A minor-allele-frequency threshold of 0.01 was specified for frequency-based filtering. The HapMap3-matched summary-statistic files were used as LDSC regression inputs.

PWAS inputs were prepared separately from GRCh38/hg38-based protein GWAS data, as described below.

### SNP-based heritability and genetic correlation

Univariate LDSC was performed for all 36 traits using the HapMap3 SNP set and European-ancestry LD-score and regression-weight files. Observed-scale heritability (h_obs_^2^ ), its standard error, the LDSC intercept, intercept standard error and mean χ² statistic were extracted. The heritability *Z* score was calculated as 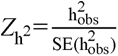. When SE(h_obs_^2^) > 0, the descriptive one-sided upper-tail Wald *P* value was calculated as 1 − Φ(Z_h_²), where Φ is the standard normal cumulative distribution function. This *P* value was not used as the primary trait-selection criterion.

Four traits with negative observed-scale heritability estimates (amoebiasis, rheumatic valve diseases, acute hepatitis A and acute peritonitis) and three traits with positive but near-zero estimates (cholera, enterobiasis and cholangitis) were classified as unstable and excluded from prioritisation, leaving 29 traits (Supplementary Data 1).

Positive near-zero estimates were classified using the study’s extraction rule. The remaining traits were screened using Z_h_² > 4 and 0 < h_obs_^2^ < 1; 16 traits met the final high-quality criteria.

For binary traits, observed-scale heritability was converted to the liability scale using population prevalence (K) and sample prevalence (*P*) [31], Where *P=N_c_ases*/(*N*_cases_+*N*_controls_). Population prevalence was taken from the external prevalence table. With t = Φ ¹(1 − K) and φ(t) denoting the standard normal density at t, liability-scale heritability was calculated as h²_liability_ =h²_obs_ ×K²(1−K)²/[φ(t)²P(1−P)]. The standard error was multiplied by the same conversion factor. Prevalence values were treated as fixed, and their uncertainty was not propagated.

Bivariate LDSC was used to evaluate all 320 possible combinations of the 16 digestive infectious disease traits and 20 cardiovascular traits. The --rg option was used with hg19-compatible munged summary statistics and European-ancestry LD-score and regression-weight files. Genetic correlation was calculated as 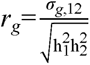, where σ ,12 is the cross-trait genetic covariance and h ² and h ² are the trait-specific SNP heritabilities. Models used free intercepts; neither phenotype-specific intercepts nor the cross-trait intercept were constrained. Because source-study sample overlap was not formally quantified, the cross-trait intercept was not fixed at zero. The sign of each genetic correlation was retained. Two-sided LDSC P values were Bonferroni-corrected across 320 pairs (*P* < 0.05/320 = 1.56 × 10 ). A double-pass quality filter required both traits in a pair to meet the univariate criteria. Thirty-nine pairs were carried forward (Supplementary Data 2).

SUPERGNOVA [32] was used to estimate local genetic covariance for the 39 retained pairs across 1,703 approximately LD-independent hg19 regions defined using the LDetect/Fourier partition [33] and European-ancestry 1000 Genomes Project Phase 3 reference data [34]. Local genetic covariance was the primary local measure. Bonferroni correction was applied within each trait pair across 1,703 blocks (*P* ≤ 0.05/1,703 = 2.94 × 10 ). Local regions with *P* < 0.05 were retained for exploratory description (Supplementary Data 3).

### MTAG and cross-trait sensitivity analyses

MTAG version 1.0.8 was applied to each of the 39 retained trait pairs after allele harmonisation. Trait 1 represented the digestive infectious disease phenotype and Trait 2 the cardiovascular phenotype. The --fdr option was enabled; the minimum minor-allele frequency was 0.001 and the median *Z*-score cutoff was 5.0.

Trait-specific MTAG effect estimates, standard errors, *P* values, mean χ² statistics and max-FDR estimates were recorded. Mean χ², max-FDR and LDSC intercepts calculated from MTAG-derived summary statistics were used jointly as diagnostic measures; no single uniform calibration cutoff was used to exclude all pairs (Supplementary Data 5).

After allele alignment, candidate pleiotropic SNPs were required to meet MTAG *P* < 5 × 10 and single-trait GWAS *P* < 0.001 for both component traits. Independent lead variants were selected by LD clumping in PLINK 1.9 [35] using the European-ancestry 1000 Genomes reference panel, with index-SNP *P* < 5 × 10 , secondary-SNP *P* < 1 × 10 , r² < 0.001 and a 500-kb window. Trait-pair-specific records were retained separately when a SNP appeared in more than one pair (Supplementary Data 4 and 6).

To assess the robustness of the prioritised cross-trait associations, we applied four complementary methods: pleiotropic analysis under a composite null hypothesis with adjustment for cross-trait correlation (PLACO+) [36], cross-phenotype meta-analysis (CPMA) [37], genetic analysis incorporating pleiotropy and annotation (GPA) [38], and cross-phenotype association analysis using the heterogeneous-effect statistic (CPASSOC-SHet) [39]. These analyses evaluated eight candidate trait-pair-variant records, representing four unique SNPs across five IID-cardiovascular disease pairs. The corresponding single-trait GWAS summary statistics were matched by SNP identifier and harmonised to the same effect allele, retaining 1,166,018-1,166,100 SNPs per pair. The MTAG *P* values reported in Supplementary Data 9 were taken from the IID-specific MTAG output for each pair, consistent with Supplementary Data 6.

PLACO+ tests whether a genetic variant is associated with both traits by considering a composite null hypothesis comprising association with neither trait or with only one trait. It combines the two single-trait *Z* statistics and accounts for their correlation, including correlation arising from overlapping samples or shared controls. We used the placo.plus function in the PLACO v0.2.0 R script. Genome-wide summary statistics were used to estimate the background *Z*-statistic variances and cross-trait correlation. For this parameter-estimation step, SNPs with *Z*² > 80 in either trait were excluded. Variances were estimated after excluding SNPs with *P* < 1 × 10 in both traits, whereas the Pearson correlation was estimated from SNPs with *P* ≥ 1 × 10 in both traits. Numerical integration used the default absolute tolerance, approximately 3 × 10 ¹³. Candidate SNP statistics were retrieved from the complete harmonised datasets; the *Z*² filter was applied only during background parameter estimation.

CPMA evaluates whether the distribution of association *P* values for a SNP across traits departs from the expectation under the null hypothesis. Under the null, *P* values follow a uniform distribution, and their negative natural logarithms follow an exponential distribution with a rate of 1. We implemented CPMA using study-specific R code that compared this null rate with the maximum-likelihood rate estimated from the two single-trait *P* values. The likelihood-ratio statistic was evaluated using a chi-square approximation with one degree of freedom. Because CPMA uses *P* values, its statistic does not encode the direction of allelic effects. No additional calibration for cross-trait dependence was applied to this two-trait implementation, and its results were interpreted as exploratory.

GPA was fitted separately to the genome-wide two-trait *P*-value matrix for each pair without functional annotation data. The model comprised four latent association states: association with neither trait (M00), IID alone (M10), the cardiovascular trait alone (M01), or both traits (M11). *P* values under null and non-null associations were modelled using uniform and beta distributions, respectively. Model parameters were estimated using the expectation-maximisation algorithm. For the shared-association pattern “11”, SNP-specific local FDR values were extracted using the fdr function and used to assess support for association with both traits.

CPASSOC-SHet combines single-trait GWAS *Z* statistics using sample-size weights while allowing heterogeneous genetic effects across traits. Cross-trait correlation is incorporated into the analysis to accommodate dependence between the input statistics. For each pair, we estimated the correlation matrix using SNPs with |*Z*| < 1.96 in both traits and supplied the median SNP-level sample size for each trait. The SHet statistic was calculated using the SHet function. Its null distribution was approximated by a shifted gamma distribution, with shape, scale and location parameters estimated using 10,000 simulated null replicates through the EstimateGamma function. Association *P* values were obtained from the upper tail of the fitted distribution.

Method-specific support was defined as *P* ≤ 5 × 10 for PLACO+, CPMA and CPASSOC-SHet, and local FDR < 0.20 for GPA. Support was recorded separately for each method in Supplementary Data 9. Single-trait GWAS and IID-specific MTAG association *P* values were two-sided. These analyses assessed consistency across statistical methods using the same underlying GWAS data and did not constitute replication in independent samples.

### Fine-mapping and Bayesian colocalisation

Fine-mapping and colocalisation were restricted to 12 independent lead-locus records across seven IID-cardiovascular disease pairs. Each trait-pair-lead-variant combination was treated as a separate record. For each lead SNP, variants within ±500 kb were evaluated. Fine-mapping was performed using the official FM-summary getCredible.r script obtained from the authors’ GitHub repository [40]. After allele harmonisation, cross-trait *Z* statistics were calculated from trait-specific MTAG *Z* scores using the sample-size-weighted Stouffer method, 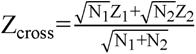; For fine-mapping, variants with discordant directions of the harmonised MTAG effect estimates were assigned a cross-trait *P* value of 1. The FM-summary implementation retained variants with LD r² > 0.4 to the lead SNP, converted association *P* values to log-evidence scores, normalised these scores to posterior probabilities and formed a 99% credible set. Under the study-defined fine-mapping support criterion, a lead SNP was retained if its normalised fine-mapping probability (probNorm) was ≥ 0.05 or it had the highest probNorm in the locus (Supplementary Data 7).

Bayesian colocalisation was performed in the same ±500-kb regions using coloc.abf in coloc v5.2.3. The primary analysis used harmonised trait-specific MTAG summary statistics. For binary traits, the case-control model used the trait-specific case proportion, sample size, effect estimate, effect-estimate variance and regional allele frequency. The coloc.abf model allowed at most one causal variant per trait in a region and used default priors (p = 1 × 10 , p = 1 × 10 and p = 1 × 10 ). The single-causal-variant assumption may limit colocalisation inference in regions with allelic heterogeneity [41]. Strong evidence of a shared signal was defined as PPH4 ≥ 0.80. Final candidate variants were required to meet the MTAG, LD-lead, FM-summary and colocalization criteria (Supplementary Data 7 and 8).

### Gene and functional prioritisation

For each retained pair, MAGMA v1.10 gene-based analysis was performed separately using the two trait-specific MTAG summary-statistic datasets, default parameters and the SNP-wise mean model [42]. LD was estimated using the European-ancestry 1000 Genomes reference panel, and SNP-to-gene mapping used an hg19-compatible annotation file. Bonferroni correction was applied separately within each component analysis using a fixed denominator of 19,248 genes (unadjusted *P* ≤ 0.05/19,248, approximately 2.60 × 10 ). The union of genes reaching Bonferroni-adjusted *P* ≤ 0.05 in either component analysis was retained for exploratory gene prioritisation. For display in Supplementary Data 10, the smaller unadjusted *P* value and the *Z* statistic from the same component analysis were reported for each gene-trait-pair record. MAGMA *Z* statistics indicate association strength rather than allelic-effect direction. Candidate pleiotropic genes were prioritised by intersecting this gene union with annotated genes on the same chromosome whose intervals overlapped a ±500-kb window around a candidate variant from the corresponding pair (Supplementary Data 8 and 14).

Gene-set enrichment [43] was conducted separately for each prioritised pair using clusterProfiler v4.18.4 [44, 45], with gene sets from GO [46] and KEGG. MAGMA genes were ranked by gene-level *Z* score; duplicate identifiers were resolved by averaging their *Z* statistics. Gene-set sizes were restricted to 10-500. Significant positive enrichment was defined as Bonferroni-adjusted P ≤ 0.05 together with NES > 2.0(Supplementary Data 11). Using MAGMA-identified pleiotropic risk genes meeting the gene-level Bonferroni-corrected *P* ≤ 0.05 threshold, tissue-specific enrichment was assessed separately for each of the five prioritised trait pairs using deTS v1.0 [47] and its GTEx-derived score matrix covering 47 tissues. The tissue-specific gene proportion was set to 0.05 (top 5%). Enrichment was evaluated using the one-sided Fisher’s exact test implemented in deTS. Bonferroni correction was applied across the 47 tissues separately within each trait pair, with adjusted P ≤ 0.05 indicating significance (Supplementary Data 12).

Cell-type-specific enrichment was assessed separately for the same five prioritised trait pairs using WebCSEA [48], with the corresponding MAGMA-significant gene lists as input. WebCSEA *P* values were evaluated against a prespecified primary Bonferroni threshold of *P* < 0.05/1,355 (approximately 3.69 × 10 ) within each trait pair. Adult tissue-cell-type entries with *P* < 1 × 10 ³ were highlighted for nominal significe (Supplementary Data 13).

Transcriptome-wide association analysis used the FUSION framework [49] with GTEx v8 expression-prediction weights [50] from 49 European-ancestry tissues and 1000 Genomes European LD covariance matrices. Candidate genes were tested using cis variants within ±500 kb; Benjamini-Hochberg correction [51] was applied across the tested gene-tissue-trait-pair-trait-side combinations, with FDR-adjusted *P* ≤ 0.05 considered significant (Supplementary Data 15).

Protein-wide association analysis used the local FUSION/PWAS implementation and ARIC plasma-protein prediction models for 1,348 cis-heritable proteins derived from 7,213 European American participants [52]. PWAS summary statistics were prepared from the GRCh38 protein GWAS data using the corresponding GRCh38 model file and the EUR LD reference; the MHC region was excluded from merged results. A single Benjamini-Hochberg correction was applied across all candidate-gene PWAS tests, with FDR-adjusted *P* ≤ 0.05 considered significant (Supplementary Data 16). Significant proteins were annotated against the Therapeutic Target Database [53] and assigned druggability tiers using the stated gene-symbol mapping [54].

### Bidirectional Mendelian randomisation

Bidirectional two-sample MR was performed for five IID-cardiovascular disease pairs carrying candidate pleiotropic variants: IID with cardiac arrhythmias, atrial fibrillation and flutter, myocardial infarction, ischaemic heart disease and venous thromboembolism (Supplementary Data 8). MR inference relies on instrument relevance, independence from confounders and the absence of effects on the outcome except through the exposure [55]. Summary statistics were from FinnGen release 10 [56], included European-ancestry participants of both sexes and used GRCh38 coordinates (Supplementary Data 17). Genetic instruments were selected at *P* ≤ 1 × 10 and LD clumping used r² = 0.001 and a 10-Mb window. Instruments with F > 10 were retained (Supplementary Data 19). Exposure and outcome alleles were harmonised, and SNPs with missing, incompatible or unresolved allele orientation were removed. An additional outcome-association filter retained SNPs with outcome GWAS *P* ≥ 0.01.

The inverse-variance-weighted (IVW) estimate [57] was the primary MR result. Fixed-effect IVW was prioritised when Cochran’s Q test did not indicate heterogeneity (*P* > 0.05); multiplicative random-effects IVW was reported when heterogeneity was detected. MR-Egger [58], weighted-median [59], weighted-mode [60] and MR-RAPS [61] estimates were used as complementary analyses. Effect estimates were reported as β and, for binary outcomes, converted to odds ratios as OR = exp(β), with 95% confidence intervals calculated as exp(β ± 1.96 × SE) (Supplementary Data 18). Heterogeneity and directional horizontal pleiotropy were assessed using Cochran’s Q and the MR-Egger intercept, respectively. RadialMR [62] was used to identify heterogeneity-based outliers, and the MR-PRESSO global test [63] was used to assess horizontal pleiotropy. Separate sensitivity analyses excluded radial MR outliers, all palindromic SNPs, or SNPs associated with potential confounding phenotypes; each filtering analysis started from the harmonised main-analysis instrument set. Confounder-associated variants were identified using the GWAS Catalog [64] for smoking, alcohol use, physical activity, body fat or adiposity-related traits, education and diabetes (Supplementary Data 19). Leave-one-out analyses used the radial-MR-filtered instrument sets with fixed-effect IVW in the forward direction and the harmonised main-analysis instrument sets with the default multiplicative random-effects IVW procedure, including correction for underdispersion, in the reverse direction (Supplementary Fig. S10). Association P values were two-sided. Benjamini-Hochberg correction was applied separately across the five forward and five reverse primary IVW tests; FDR-adjusted P ≤ 0.05 indicated significance. Complementary and sensitivity-analysis association P values were reported without multiple-testing correction (Supplementary Data 18).

### Gut microbiome mediation analysis

Two-step MR mediation analyses evaluated the prespecified pathway from a gut microbial feature through IID to a cardiovascular outcome [65]. The five outcomes were cardiac arrhythmias, venous thromboembolism, myocardial infarction, atrial fibrillation and flutter, and ischaemic heart disease. Microbial instruments were obtained from three independent resources: the German microbiome GWAS (430 features; 8,956 participants) [66], the Dutch Microbiome Project (DMP) cohort (412 features; 7,738 participants) [67] and the MiBioGen consortium (24 cohorts; 18,340 participants; 211 taxonomic features) [17].

For each microbial feature, instruments were selected at *P* < 1 × 10 and clumped at r² < 0.001. Allele harmonisation and instrument quality control followed the disease-level MR procedures. The IVW estimate for the microbial-feature-to-IID association was denoted alpha, and the corresponding primary IID-to-cardiovascular MR estimate was denoted beta. The total microbial-feature-to-cardiovascular effect was estimated separately using IVW (Supplementary Data 18 and 20). The indirect effect was alpha × beta, the direct effect was the total effect minus the indirect effect, and the mediated proportion was the indirect effect divided by the total effect (Supplementary Data 21).

Standard errors and 95% confidence intervals for the indirect effect and mediated proportion were estimated using the delta method. Unavailable covariance terms were assumed to be zero. Direct-effect confidence intervals were approximated from the standard errors of the total and indirect effects under an independence assumption (Supplementary Data 21). Supplementary Data 20 reports two-sided *P* values for the component microbial-feature MR associations. Indirect-effect *P* values were also two-sided. The selected mediation patterns were interpreted as exploratory. Directionally concordant positive patterns were prioritised for the main text, while protective and directionally inconsistent patterns were retained in Supplementary Data 21. These summary-statistics-based analyses were interpreted as putative genetically mediated patterns rather than experimental evidence of biological mediation.

### Statistical analysis, software and reproducibility

*P* values were interpreted according to the test implemented by each method. GWAS and MR association *P* values were two-sided; descriptive heritability Wald *P* values used a one-sided upper tail, and deTS used a one-sided Fisher’s exact enrichment test. Heterogeneity, MR-PRESSO and other method-specific *P* values were reported as returned by the corresponding software. Multiple-testing procedures were applied within the prespecified test families described above. Trait-level heritability screening did not receive an additional multiple-testing correction. Software included LDSC v1.0.0 for univariate analyses and v1.0.1 for cross-trait analysis, the official SUPERGNOVA implementation, MTAG v1.0.8, FM-summary(https://github.com/hailianghuang/FM-summary), coloc v5.2.3, PLACO v0.2.0, GPA v1.22.0, MAGMA v1.10, clusterProfiler v4.18.4, org. Hs.eg.db v3.22.0, deTS v1.0, WebCSEA (accessed 10 April 2026), the FUSION/TWAS framework v1.4.2, TwoSampleMR v0.6.24 [68], mr.raps v0.4.2 and RadialMR v1.2.1. Analyses were conducted using R v4.5.2 and Python v3.13.9. CPMA used study-specific R code based on the method of Cotsapas et al.; CPASSOC-SHet used the local EstimateGamma and SHet source functions following Zhu et al., with no numbered release encoded in the source files. The local PWAS implementation also had no separate numbered release recorded.

All analysed genetic data were publicly available, de-identified GWAS summary statistics; no individual-level participant genotype or phenotype data were analysed. Under the applicable institutional policy, no additional ethics approval was required. Analysis scripts and parameter files will be deposited in a public repository, with the accession or DOI added to the final manuscript.

## Results

### 1. Shared genetic architecture between digestive infectious diseases and cardiovascular diseases

#### SNP-based heritability and trait prioritisation

In the European-ancestry population, we first estimated SNP-based heritability for 16 digestive infectious disease traits and 20 cardiovascular traits included in the study (Supplementary Data 1). Raw univariate linkage disequilibrium score regression (LDSC) outputs provided observed-scale heritability estimates for all 36 curated traits. These estimates ranged from negative or near-zero values to 9.2%, indicating substantial variation in the strength of trait-level genetic signals. Seven traits yielded negative or near-zero heritability estimates and were therefore regarded as unstable for trait prioritisation, rather than as having missing heritability estimates. Exclusion of these traits left 29 phenotypes with usable observed-scale heritability estimates.

Among the retained traits, observed-scale heritability ranged from 0.1% for enterocolitis due to *Clostridium difficile* to 1.2% for intestinal infectious diseases (IID) among the digestive infectious disease traits, and from 0.1% for pericarditis to 9.2% for atrial fibrillation and flutter among the cardiovascular traits. We subsequently applied the predefined quality-control criteria of heritability *Z* score >4 and h² <1, retaining 16 high-quality traits for downstream analyses. Because all eligible traits had h² estimates below 1, this filtering step was effectively determined by the *Z* score >4 criterion. The final set comprised three digestive infectious disease traits and 13 cardiovascular traits (Supplementary Data 1). Liability-scale heritability ranged from 3.5% for IID to 13.1% for viral hepatitis among the digestive infectious disease traits, and from 4.9% for cerebrovascular disease to 22.7% for peripheral angiopathy among the cardiovascular traits (Supplementary Data 1; Fig. 2a, panel A). These results indicated measurable heritable components across the prioritised phenotypes and supported their inclusion in subsequent cross-trait analyses.

**Fig. 2.**
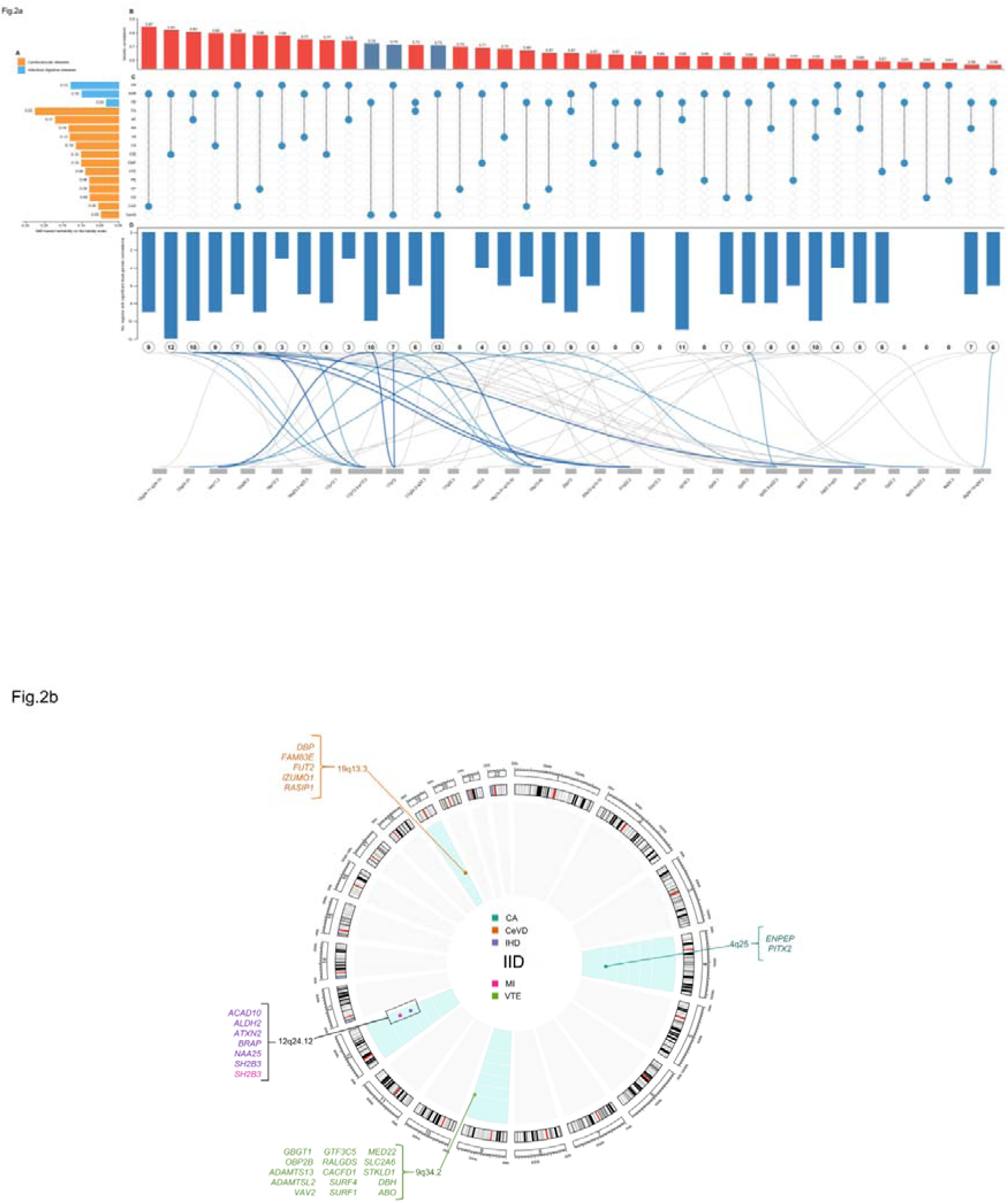
Heritability and shared genetic architecture between digestive infectious diseases and cardiovascular traits. **a** Heritability and genetic sharing between digestive infectious diseases and cardiovascular traits. **(A)** Liability-scale SNP heritability (h^2^ ) estimated using LDSC for 16 prioritised traits. Blue and orange bars denote digestive infectious and cardiovascular traits, respectively. **(B)** Global genetic correlations for 39 quality-filtered trait pairs passing Bonferroni correction across 320 tests (*P* < 0.05/320). Bars show (|r_g_|); red indicates positive correlations and blue negative correlations. **(C)** Trait-pair membership matrix. Vertically connected filled circles identify each pair; columns align across B - D. **(D)** SUPERGNOVA local genetic covariance across 1,703 approximately LD-independent blocks. Downward bars and circled values count regions with unadjusted *P* < 0.05; curves show the *P* < 0.01 subset and link pairs to cytoband intervals. Dark blue curves denote the eight lowest *P* values, light blue the next 12, and grey the remainder. Grey tile widths increase with mapped association counts. Local signals are exploratory; none passed within-pair Bonferroni correction (*P* ≤ 0.05/1,703). Underlying data for panels A, B-C and D are provided in Supplementary Data 1, 2 and 3, respectively. **b** Genomic distribution of selected IID-centred MTAG signals and regional gene annotations. Outer rings show chromosome positions (Mb) and cytogenetic bands; pale blue sectors highlight chromosomes containing displayed signals. Coloured dots mark signal positions, and dot and gene-label colours denote cardiovascular traits. The black cytoband label and dotted rectangle highlight a region shared by multiple trait pairs. This overview precedes the final fine-mapping and colocalisation filter and includes the broader cerebrovascular signal. Regional gene labels are annotations rather than the final set of 24 candidate genes in Supplementary Data 14; the final candidate genes are shown in Supplementary Fig. S6. Lead-variant records are provided in Supplementary Data 6. Abbreviations: AA, aortic aneurysm; AAR, abscess of anal and rectal regions; AF, atrial fibrillation and flutter; CA, cardiac arrhythmias; CAD, coronary artery disease; CD, conduction disorders; CeVD, cerebrovascular disease; CMP, cardiomyopathy; HF, heart failure; IID, intestinal infectious diseases; IHD, ischaemic heart disease; LD, linkage disequilibrium; LDSC, linkage disequilibrium score regression; MI, myocardial infarction; PA, peripheral angiopathy; PE, pulmonary embolism; SNP, single-nucleotide polymorphism; VH, viral hepatitis; VTE, venous thromboembolism; (r_g_), genetic correlation.

#### Global genetic correlations

We next evaluated global genetic correlations across the 16 prioritised traits. Cross-trait LDSC identified significant genetic correlations for all 39 digestive infection-cardiovascular disease trait pairs after Bonferroni correction (Supplementary Data 2; Fig. 2a, panels B and C). Genetic correlation coefficients ranged from −0.7447 for IID-cerebrovascular disease (*P*=1.36×10 ²) to 0.8662 for abscess of the anal and rectal regions-coronary artery disease (*P*=2.11×10 ³ ).

Of the 39 significant trait pairs, 36 showed positive genetic correlations and three showed negative correlations. All three negative correlations involved cerebrovascular disease and were observed for IID-cerebrovascular disease, viral hepatitis-cerebrovascular disease, and abscess of the anal and rectal regions-cerebrovascular disease. By contrast, broad positive genetic correlations were observed between digestive infectious disease traits and cardiovascular phenotypes related to cardiac rhythm, ischaemia, thrombosis, coronary disease, and heart failure. These findings demonstrated widespread but directionally structured genetic sharing between digestive infectious diseases and cardiovascular diseases. Univariate LDSC metrics indicated generally acceptable calibration of the underlying GWAS summary statistics. For digestive infectious disease traits, mean χ² values ranged from 1.0563 to 1.1300 and LDSC intercepts ranged from 1.0108 to 1.0321.

For cardiovascular traits, mean χ² values ranged from 1.0438 to 1.6092, with LDSC intercepts ranging from 1.0005 to 1.1715 (Supplementary Data 2). Most intercepts were close to 1, although higher values were observed for some cardiovascular phenotypes. Thus, the metrics did not indicate pervasive inflation across the analysed GWAS datasets, although residual inflation could not be excluded for individual cardiovascular traits.

#### Local genetic correlations

At the local level, we used SUPERGNOVA to evaluate genetic covariance across 1,703 approximately LD-independent genomic blocks. No local covariance signal remained significant after Bonferroni correction. Nevertheless, multiple nominally significant local covariance signals were observed at *P*<0.05, and the number of such regions varied substantially among trait pairs, suggesting heterogeneity in the distribution of local genetic sharing (Supplementary Data 3). For the integrated visualisation, regional counts are shown at unadjusted *P* < 0.05, whereas pair-cytoband links are restricted to *P* < 0.01 (Fig. 2a, panel D).

Several cytoband intervals recurred among the nominal signals across multiple trait pairs, including 3p22.3-p22.2, 2p25.3, 9q34.13-q34.2, 19q13.31-q13.32, and 17q24.2-q24.3. Cerebrovascular disease-related pairs showed recurrent nominal signals at 17q12, 14q11.2, 19q13.42, and 8p23.3-p23.2, whereas thromboembolic and rhythm-related outcomes more frequently involved 9q34.13-q34.2, 21q22.2, and 3p22.3-p22.2. IID-related pairs contained between five and 11 nominally significant local regions across different cardiovascular outcomes (Supplementary Data 3).

Because none of these local signals survived multiple-testing correction, they were considered exploratory rather than definitive evidence of local pleiotropy. Overall, the distribution of nominal signals was consistent with a potentially polyregional shared architecture, whereas robust local genetic sharing at individual genomic blocks was not established.

Given its stable heritability and significant global genetic correlations with all 13 prioritised cardiovascular traits, IID was selected as the principal digestive infectious disease phenotype for subsequent cross-trait mapping and functional analyses (Supplementary Data 1 and 2).

### 2. Cross-trait meta-analysis identifies IID-centred pleiotropic loci

Using the 39 digestive infection-cardiovascular disease trait pairs with significant global genetic correlations, we performed multi-trait analysis of GWAS (MTAG) to identify pleiotropic genetic variants. MTAG-derived calibration and inflation metrics varied across trait pairs, indicating that the performance of the cross-trait analysis was not uniform across all digestive infectious disease phenotypes. Nevertheless, variant-level pleiotropic signals were strongly concentrated in IID-related trait pairs. Across all 39 trait pairs, we identified 41 trait-pair-specific pleiotropic SNP records and 14 independent lead variants, representing eight unique lead SNPs overall (Supplementary Data 4-6; Supplementary Fig. S1). IID-related pairs accounted for 39 candidate pleiotropic SNP and 12 independent lead variants, representing seven unique lead SNPs. These signals were primarily localised to 4q25, 9q34.2, 12q24.12-q24.13, and 19q13.33 and involved cardiovascular phenotypes related to cardiac rhythm, thrombosis, ischaemia, coronary disease, and cerebrovascular disease (Fig. 2b). Viral hepatitis-related pairs showed only limited evidence of pleiotropy, whereas no pleiotropic variants were identified for pairs involving abscess of the anal and rectal regions.

IID-related pairs also generally showed more favourable trait-1 MTAG maxFDR estimates than pairs involving the other digestive infectious disease phenotypes, indicating a lower estimated susceptibility to false discoveries arising from violations of the MTAG homogeneity assumptions. Accordingly, although Supplementary Data 5 summarises all 39 analysed trait pairs, subsequent variant-level analyses focused primarily on IID-related signals.

MTAG-derived calibration metrics did not indicate widespread technical inflation. Across the analysed trait pairs, mean χ² values ranged from 1.042 to 1.096 for the digestive infectious disease trait side and from 1.041 to 1.407 for the cardiovascular trait side. The corresponding LDSC intercepts calculated from the MTAG-derived summary statistics ranged from 0.8478 to 1.0000 for the digestive infectious disease traits and from 0.9807 to 1.0047 for the cardiovascular traits (Supplementary Data 5). Cardiovascular-trait intercepts remained tightly clustered around 1, whereas intercepts below 1 for several digestive infectious disease traits were more consistent with conservative or model-dependent calibration than with excessive inflation.

Collectively, these findings identified IID as the predominant digestive infectious disease phenotype contributing to pleiotropic genetic sharing with cardiovascular diseases and motivated the restriction of subsequent fine-mapping and colocalization analyses to IID-centred loci.

### 3. Fine-mapping and colocalization identify candidate pleiotropic variants

We next applied FM-summary fine-mapping and Bayesian colocalization to refine the IID-related pleiotropic loci identified through MTAG. In total, 12 trait-pair-specific independent pleiotropic loci across seven IID-cardiovascular disease pairs were taken forward for variant-level analysis (Supplementary Data 6).

Under the predefined fine-mapping criteria, eight of the 12 lead-SNP records were retained. Colocalization analysis provided evidence for a shared causal signal at 11 of the 12 loci, defined as a posterior probability for hypothesis 4 (PPH4) ≥ 0.8. The only locus without colocalization evidence was the rs579459 locus in the IID-cerebrovascular disease pair, for which PPH4 was 0.0016 (Supplementary Data 7).

Integration of the fine-mapping and colocalization results yielded eight candidate trait-pair-variant records, representing four unique candidate pleiotropic variants: rs486632, rs579459, rs653178, and rs17630235 (Supplementary Data 8; Supplementary Fig. S2 ). These variants were concentrated at 4q25, 9q34.2, and 12q24.12-q24.13.

The rs579459 variant at 9q34.2 was retained in three IID (intestinal infectious diseases)-related cardiovascular trait pairs: cardiac arrhythmias, venous thromboembolism, and atrial fibrillation and flutter. The rs486632 variant at 4q25 was shared by the IID-cardiac arrhythmias and IID-atrial fibrillation and flutter pairs. At 12q24, rs653178 was shared by the IID-myocardial infarction and IID-ischaemic heart disease pairs, whereas rs17630235 represented an additional candidate variant for IID-ischaemic heart disease. The locus-to-trait-pair relationships are summarised in Supplementary Fig. S3.

Several additional loci showed strong colocalization evidence but were not retained as final candidate variants because their independent lead SNPs did not satisfy the fine-mapping criteria. These included rs11066188 and rs3184504 in the IID-coronary artery disease pair, with PPH4 values of 0.9976 and 0.9980, respectively, and rs281380 in the IID-cerebrovascular disease pair, with PPH4=0.9957. Conversely, the rs579459 locus in the IID-cerebrovascular disease pair was supported by the upstream pleiotropy analysis but not by colocalization (Supplementary Data 7).

All eight candidate trait-pair-variant records showed concordant support across the complementary PLACO+, CPMA, CPASSOC-SHet and GPA analyses. All records met the method-specific support thresholds of *P* ≤ 5 × 10 for PLACO+, CPMA and CPASSOC-SHet and GPA local FDR < 0.20 (Supplementary Data 9; Supplementary Figs. S4 and S5).

These analyses refined the broad trait- and locus-level genetic overlap between IID and cardiovascular diseases to a limited set of recurrent, high-confidence candidate pleiotropic variants.

### 4. Gene-level prioritisation and functional characterisation MAGMA gene-based analysis

We performed MAGMA gene-based association analysis using the MTAG-derived summary statistics to evaluate gene-level genetic sharing across the retained IID-cardiovascular disease pairs. Genes reaching a Bonferroni-corrected significance level of *P*_Bonferroni_ ≤ 0.05 were considered significant, and the complete results are provided in Supplementary Data 10.

MAGMA identified significant IID-related pleiotropic risk genes across the retained trait pairs, with substantial variation in the number and composition of significant genes among cardiovascular outcomes. Rhythm-related pairs showed highly concordant gene-level patterns. For IID-cardiac arrhythmias, representative significant genes included *KCND3*, *KCNN3*, *SCN5A*, *SCN10A*, *PITX2*, *ENPEP*, and *TTN*. A similar profile was observed for IID-atrial fibrillation and flutter, including *KCND3*, *KCNN3*, *SCN5A*, *SCN10A*, *PITX2*, *ENPEP*, *ANK2*, and *FAM13B*.

For IID-venous thromboembolism, significant genes were strongly related to coagulation and thrombosis and included *F11*, *ABO*, *ADAMTS13*, *CACFD*1, *SLC2A6*, *RALGDS*, and *VAV2*, consistent with the recurrent 9q34.2 signal identified at the variant level. For IID-myocardial infarction, significant genes included established coronary and atherosclerotic susceptibility genes such as *PHACTR1*, *LPA*, *LPL*, *CDKN2A*, *CDKN2B*, *ADAMTS7*, *COL4A2*, and *LIPA*. The IID-ischaemic heart disease pair included *LDLR*, *APOE*, *SMARCA4*, *PROCR*, *GGT7*, and *NCOA6* among its significant genes (Supplementary Data 10).

These results revealed outcome-specific but biologically coherent gene-level signatures related principally to cardiac electrophysiology, coagulation and thrombosis, and ischaemic vascular disease (Supplementary Data 10).

#### Prioritisation of candidate pleiotropic genes

We next intersected the MAGMA-significant genes with genes located within ±500 kb of the final candidate pleiotropic variants. This analysis yielded 30 trait-pair-SNP-gene records, corresponding to 24 unique candidate pleiotropic genes across five candidate variant records from four IID-cardiovascular disease pairs (Supplementary Data 14; Supplementary Fig. S6).

At 4q25, the rs486632 locus prioritised *PITX2* and *ENPEP* in both the IID-cardiac arrhythmias and IID-atrial fibrillation and flutter pairs, supporting this region as a recurrent electrophysiological susceptibility locus. At 9q34.2, the rs579459 locus contributed 16 candidate genes in the IID-venous thromboembolism pair, including *ABO*, *ADAMTS13*, *CACFD1*, *VAV2*, *RALGDS*, *SLC2A6*, *GTF3C5*, *ADAMTSL2*, and *GBGT1* (Supplementary Data 14; Supplementary Fig. S6 panel B). This region therefore represented the largest prioritised coagulation- and thrombosis-related gene module.

At 12q24.12-q24.13, the rs653178 and rs17630235 loci prioritised six candidate genes for IID-ischaemic heart disease: *SH2B3*, *ATXN2*, *NAA25*, *ALDH2*, *BRAP*, *and ACAD10* (Supplementary Data 14; Supplementary Fig. S6 panel B). These findings supported 12q24 as a recurrent locus connecting IID with ischaemic and vascular phenotypes.

Candidate pleiotropic genes were not recovered for every candidate variant record. In particular, although the IID-myocardial infarction pair contained a candidate pleiotropic variant in the upstream analysis, no Bonferroni-significant MAGMA gene overlapped the predefined ±500-kb window around its lead SNP. Consequently, candidate pleiotropic genes were ultimately prioritised for four of the five IID-cardiovascular disease pairs selected for downstream functional analyses (Supplementary Data 14).

#### Gene-set, tissue-specific, and cell-type-specific enrichment

Gene-set enrichment analysis was performed across the five prioritised IID-cardiovascular disease pairs. At Bonferroni-corrected *P* ≤ 0.05 and NES > 2.0, 43 trait-pair-GO term associations were identified across three pairs: 25 for IID-venous thromboembolism, 11 for IID-myocardial infarction and seven for IID-ischaemic heart disease (Supplementary Data 11; Fig. 3a). These comprised 34 biological process, four cellular component and five molecular function records. Enrichment for IID-venous thromboembolism predominantly involved coagulation, haemostasis, platelet activation and fibrinolysis. IID-myocardial infarction showed enrichment related to lipoprotein regulation, cholesterol transport and cholesterol efflux, whereas IID-ischaemic heart disease showed enrichment involving lipoprotein regulation, organisation and particle components. No KEGG pathway met the predefined enrichment criteria.

**Fig. 3.**
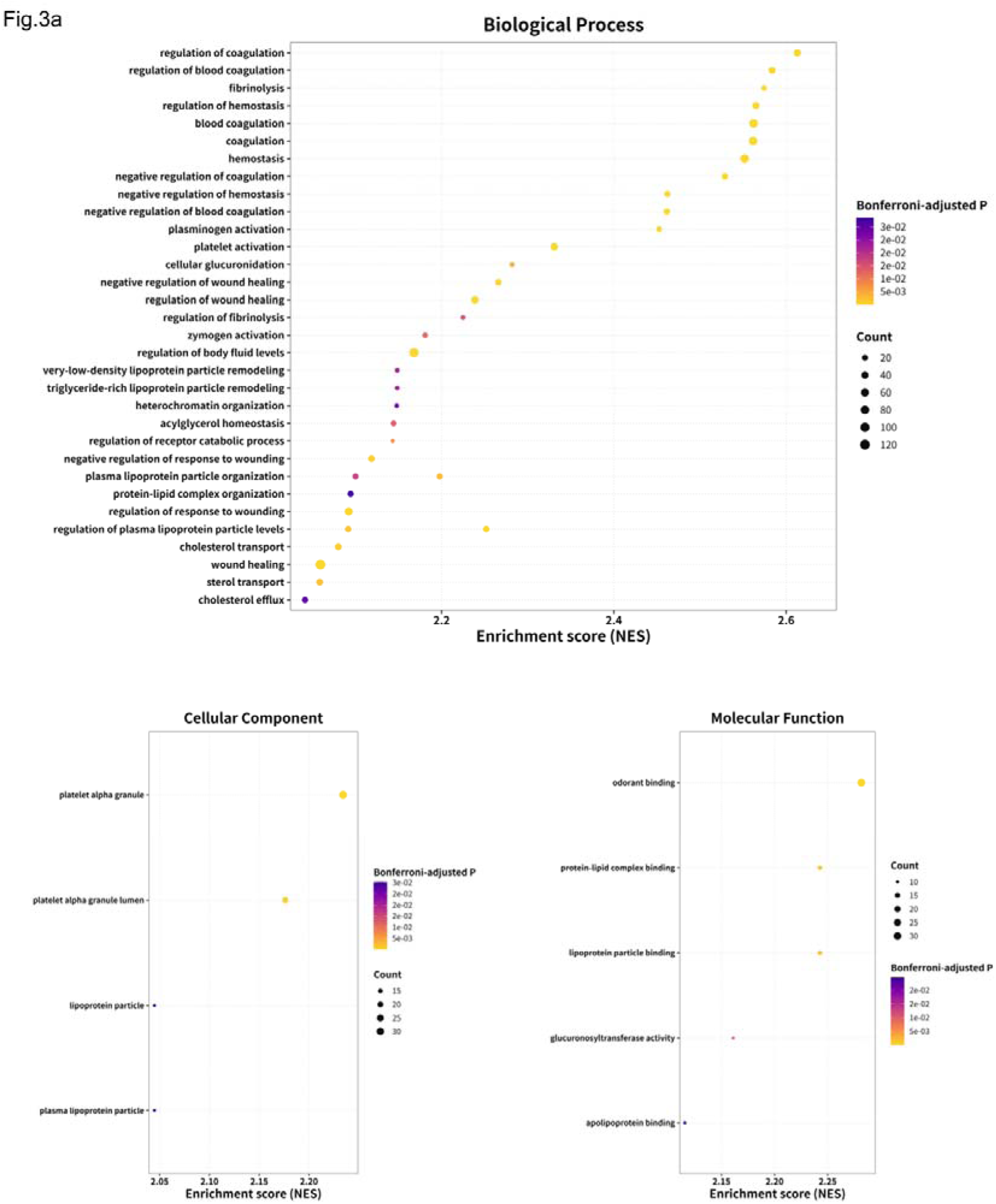
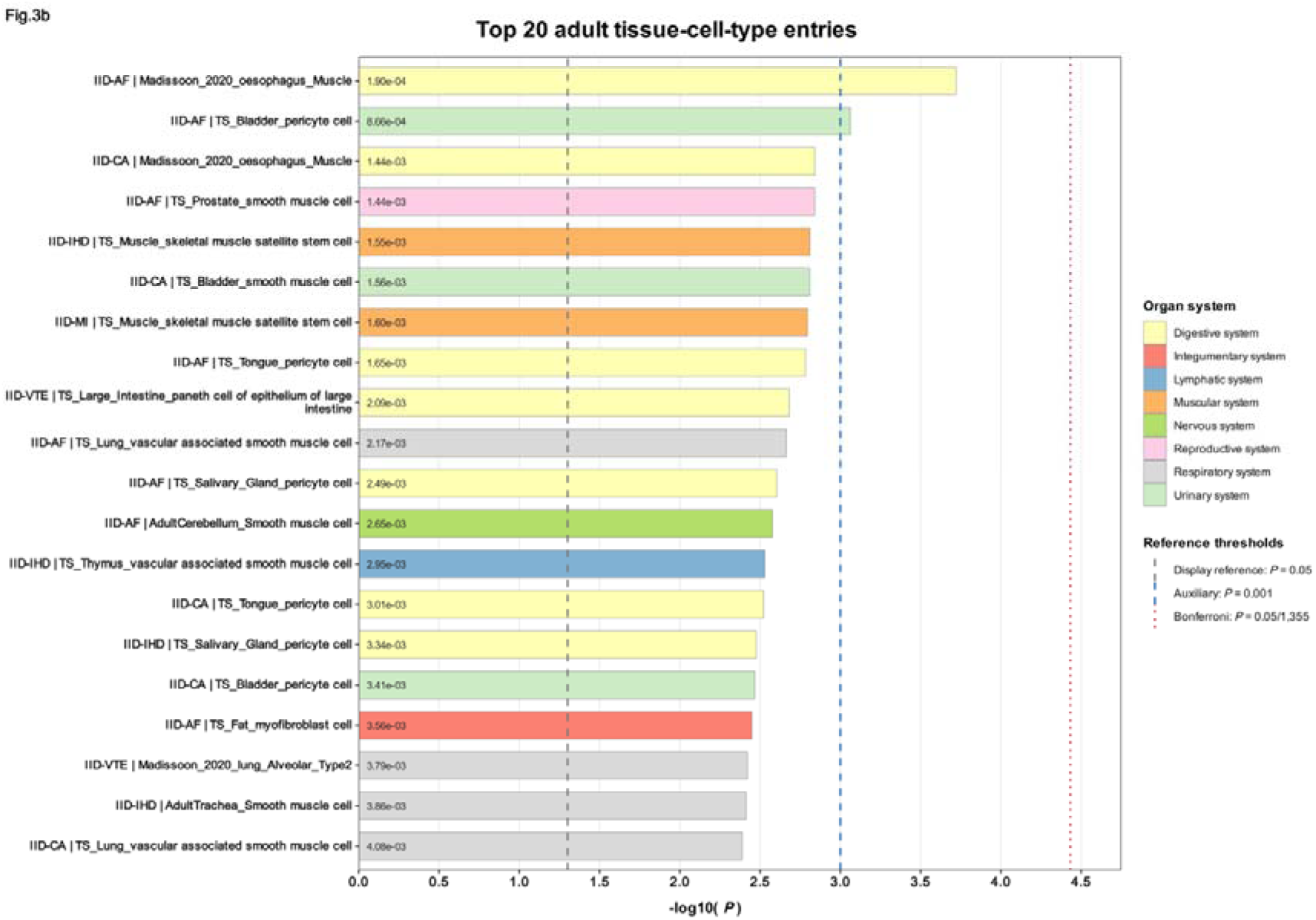
Functional characterisation of the shared genetic architecture between intestinal infectious diseases and cardiovascular traits. **a** Gene-set enrichment analysis. Bubble plots show all 43 trait-pair-GO term records in Supplementary Data 11 meeting Bonferroni-adjusted *P* ≤ 0.05 and NES > 2.0: 34 biological process records (upper), four cellular component records (lower left) and five molecular function records (lower right). Each dot represents one trait-pair-GO term record; trait-pair identities are provided in Supplementary Data 11. Horizontal position indicates NES, dot size indicates the number of unique leading-edge genes, and colour indicates the Bonferroni-adjusted *P* value (yellow, smaller *P*; purple, larger *P*). GO terms occurring in multiple trait pairs are retained as separate points. No KEGG pathway met the prespecified criteria. **b** Cell-type-specific enrichment analysis. Bars show the 20 adult tissue-cell-type entries with the smallest WebCSEA combined *P* values across the five prioritised trait pairs. Input genes were MAGMA-identified pleiotropic risk genes meeting gene-level Bonferroni-corrected *P* ≤ 0.05. Bar length indicates −log (*P*), colour denotes the organ system, and numbers within bars indicate combined *P* values. The grey dashed, blue dashed and red dotted lines mark *P* = 0.05 (display reference), *P* = 0.001 (auxiliary exploratory threshold) and *P* = 0.05/1,355 ≈ 3.69 × 10 (primary Bonferroni threshold per trait pair), respectively. Two entries, both IID-AF, reached the auxiliary threshold; none reached the primary threshold. The ranking is descriptive. Complete results are provided in Supplementary Data 13. Abbreviations: AF, atrial fibrillation and flutter; BP, biological process; CA, cardiac arrhythmias; CC, cellular component; CSEA, cell-type-specific enrichment analysis; GO, Gene Ontology; GSEA, gene-set enrichment analysis; IID, intestinal infectious diseases; IHD, ischaemic heart disease; KEGG, Kyoto Encyclopedia of Genes and Genomes; MF, molecular function; MI, myocardial infarction; NES, normalised enrichment score; VTE, venous thromboembolism.

Tissue-specific enrichment analysis identified seven significant tissue-trait-pair associations across three of the five prioritised pairs after Bonferroni correction across 47 tissues within each pair (Supplementary Data 12; Supplementary Fig. S7). IID-atrial fibrillation and flutter showed enrichment in Heart—Atrial Appendage, Heart—Left Ventricle, Artery—Tibial and Esophagus—Muscularis. IID-cardiac arrhythmias was enriched in Heart—Atrial Appendage and Heart—Left Ventricle, whereas IID-venous thromboembolism was enriched in Liver. No tissue reached the Bonferroni-corrected significance threshold for IID-myocardial infarction or IID-ischaemic heart disease.

Cell-type-specific enrichment analysis did not identify any cell type that remained significant after Bonferroni correction across the five prioritised pairs (Supplementary Data 13; Fig. S8). Under the auxiliary WebCSEA display threshold of *P* < 1×10 ³, the strongest nominal signal was observed for adult oesophageal muscle cells in the IID-atrial fibrillation and flutter pair (*Madissoon_2020_oesophagus_Muscle*; *P* = 1.90×10 ) (Supplementary Data 13). For descriptive visualisation, the 20 adult tissue-cell-type entries with the smallest WebCSEA *P* values and nominal enrichment (*P* < 0.05) are presented in (Fig. 3b).

#### Transcriptome-wide association analysis

We performed transcriptome-wide association analysis to evaluate whether the genetically predicted expression of the candidate pleiotropic genes across 49 GTEx v8 tissues was associated with the MTAG-derived IID-cardiovascular disease signals. Benjamini-Hochberg correction was applied globally across all tested gene-tissue-trait-side combinations.

Twenty-one candidate pleiotropic genes showed at least one significant association after global FDR correction (*P*_FDR_ ≤ 0.05; Supplementary Data 15 and Supplementary Fig. S9). Significant signals were distributed across blood, vascular, cardiac, gastrointestinal, adipose, neural, and cultured fibroblast tissues.

The strongest association was observed for *ABO*, located near the rs579459 locus. Genetically predicted *ABO* expression in whole blood was strongly associated with the cardiovascular trait side of the IID-venous thromboembolism pair (TWAS.*Z* = 29.82, TWAS.*P* = 1.87×10 ¹ , *P*_FDR_ = 1.20×10 ¹ ²; Supplementary Data 15).

*ENPEP* represented a prominent transcriptomic signal for the arrhythmia-related pairs. In addition, the 12q24 gene module comprising *ALDH2*, *NAA25*, *ATXN2*, *BRAP*, *SH2B3*, and *ACAD10* showed significant associations in the IID-ischaemic heart disease pair. These findings provided transcriptome-level support for several of the prioritised pleiotropic loci.

#### Protein-wide association analysis and druggability annotation

Protein-wide association analysis identified four significant protein-level associations after FDR correction (*P*_FDR_ ≤ 0.05; Supplementary Data 16). These comprised *ABO* and *OBP2B* for IID-venous thromboembolism and ENPEP for both IID-cardiac arrhythmias and IID-atrial fibrillation and flutter.

The strongest PWAS association was observed for *ABO* in IID-venous thromboembolism (PWAS.*Z* = 8.1885, PWAS.*P* = 2.64×10 ¹ , *P*_FDR_ = 3.17×10 ¹ ). *OBP2B* was also significant for the same trait pair (PWAS.*Z* = −3.5668, PWAS.*P* = 3.61×10 , *P*_FDR_ = 0.00217). *ENPEP* was significantly associated with IID-cardiac arrhythmias (PWAS.*Z* = 2.7321, PWAS.*P* = 0.00629, *P*_FDR_ = 0.0252) and IID-atrial fibrillation and flutter (PWAS.*Z* = 2.5543, PWAS.*P* = 0.0106, *P*_FDR_=0.0318) (Supplementary Data 16).

Druggability annotation classified *ENPEP* as a Tier 2 target with both small-molecule and biologic druggability support. *OBP2B* was assigned to Tier 3A and had biologic, but not small-molecule, druggability support. *ABO* did not have a matched target-drug record in the Therapeutic Target Database or an assigned druggability tier in the current annotation workflow (Supplementary Data 16).

### 5. Bidirectional Mendelian randomization supports selective causal relationships

We conducted bidirectional two-sample Mendelian randomization analyses for the five IID-cardiovascular disease pairs carrying final candidate pleiotropic variants (Supplementary Data 8). Statistical significance in the primary analysis was defined as an FDR-corrected *P* value ≤0.05 for the inverse-variance-weighted estimate.

Four significant associations were identified in the forward direction from IID to cardiovascular outcomes. Genetically predicted liability to IID was positively associated with cardiac arrhythmias (OR_IVW_ = 1.12, *P*_FDR_ = 2.33×10 ), myocardial infarction (OR_IVW_ = 1.10, *P*_FDR_ = 0.0286), atrial fibrillation and flutter (OR_IVW_ = 1.11, *P*_FDR_ = 0.0286), and ischaemic heart disease (OR_IVW_ = 1.08, *P*_FDR_ = 0.00962). No significant association was observed between IID and venous thromboembolism (Supplementary Data 18; Fig.4).

**Fig. 4.**
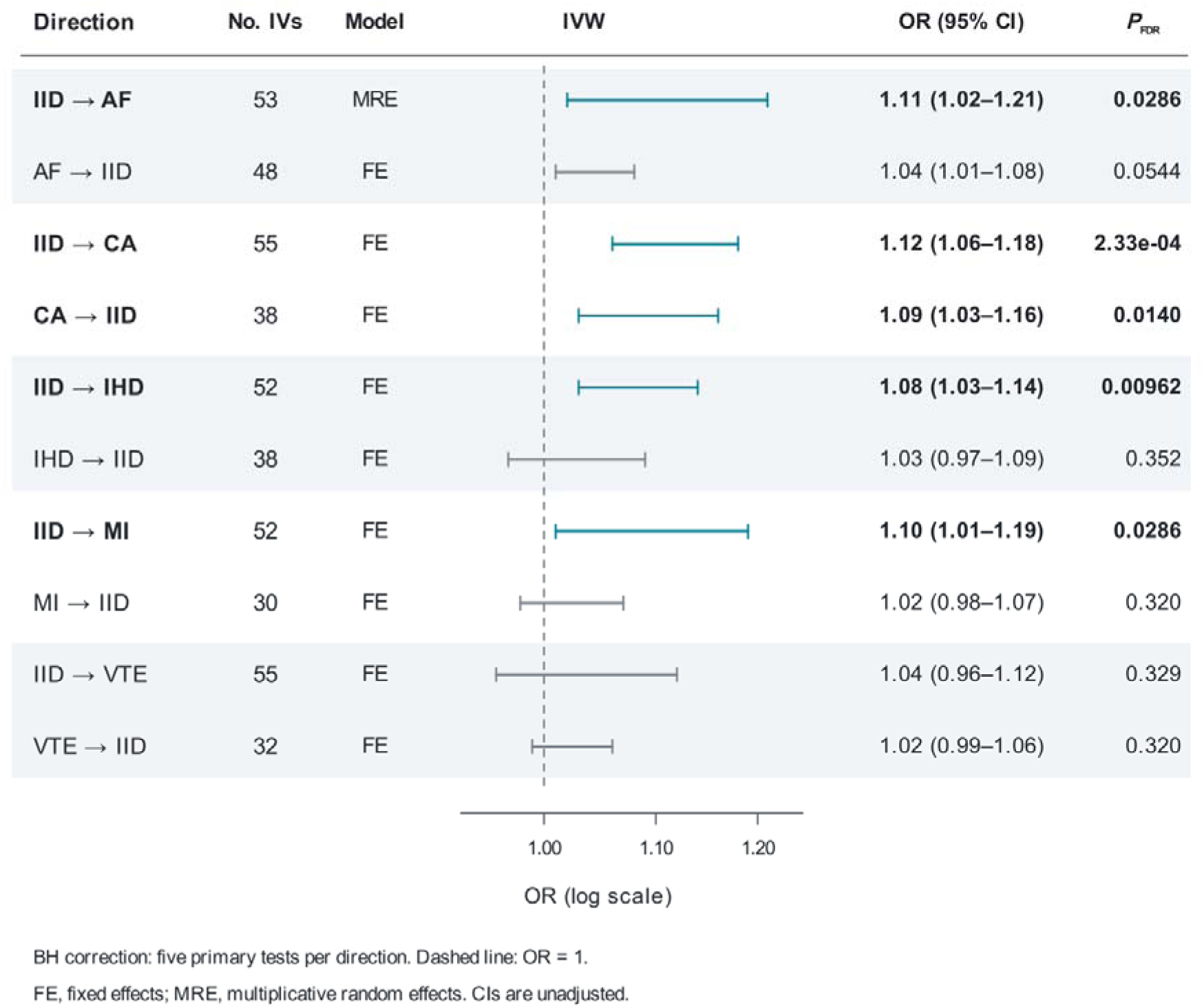
Bidirectional Mendelian randomisation estimates between intestinal infectious diseases and cardiovascular diseases. The forest plot presents primary inverse-variance-weighted (IVW) estimates for five prioritised IID-cardiovascular disease pairs, with forward and reverse analyses displayed consecutively for each pair. Horizontal lines show unadjusted 95% confidence intervals (CI); odds ratios (OR) are displayed on a logarithmic axis. The vertical dashed line indicates OR = 1. Teal intervals and bold text indicate associations reaching FDR-adjusted *P* ≤ 0.05; grey intervals indicate associations not reaching this threshold. Benjamini-Hochberg correction was applied separately across the five forward and five reverse primary IVW tests using two-sided association *P* values. Fixed-effect IVW was used when Cochran’s Q *P* > 0.05; multiplicative random-effects IVW was used otherwise. Numerical estimates and instrument characteristics are provided in Supplementary Data 18 and 19, respectively. Abbreviations: AF, atrial fibrillation and flutter; CA, cardiac arrhythmias; CI, confidence interval; FDR, false discovery rate; FE, fixed effects; IID, intestinal infectious diseases; IHD, ischaemic heart disease; IV, instrumental variable; IVW, inverse variance weighted; MI, myocardial infarction; MRE, multiplicative random effects; OR, odds ratio; VTE, venous thromboembolism.

In the reverse direction, only genetically predicted liability to cardiac arrhythmias was significantly associated with an increased risk of IID (OR_IVW_ = 1.09, *P*_FDR_ = 0.0140).

Reverse associations from myocardial infarction, atrial fibrillation and flutter, venous thromboembolism, and ischaemic heart disease to IID did not reach the predefined FDR-corrected significance threshold. Thus, IID and cardiac arrhythmias represented the only trait pair with significant evidence in both causal directions (Supplementary Data 18; Fig.4).

Several sensitivity analyses were directionally consistent with the primary findings. At an unadjusted *P* < 0.05, the associations from IID to cardiac arrhythmias and ischaemic heart disease, together with the reverse association from cardiac arrhythmias to IID, persisted across the reported radial-outlier, palindromic-variant and confounder-variant filtering analyses. For IID to ischaemic heart disease, the confounder-filtered estimate was unchanged because no instruments were removed in that filtering set. The IID-to-atrial fibrillation and flutter association showed evidence of heterogeneity in the primary analysis but remained directionally stable after radial outlier removal. By contrast, the IID-to-myocardial infarction association was attenuated and no longer reached unadjusted *P* < 0.05 after confounder-associated variants were excluded, indicating comparatively limited robustness. The IID-to-venous thromboembolism association was not statistically significant in the primary or reported filtering sensitivity analyses (Supplementary Data 18 and 19). Sensitivity-analysis *P* values were unadjusted and did not replace the FDR-adjusted primary tests. Leave-one-out analyses for both directions are presented in Supplementary Fig. S10.Leave-one-out sensitivity analyses for both directions are presented in Supplementary Fig. S10.

Overall, these results supported directionally asymmetric causal relationships between IID and selected cardiovascular diseases, with the strongest bidirectional evidence observed between IID and cardiac arrhythmias.

### 6. Gut microbiota mediation analysis identifies potential mechanistic intermediates

We integrated host genetic data with three gut microbiome GWAS resources to investigate whether microbial traits might contribute to the associations between IID and cardiovascular diseases. Focusing on directionally concordant, risk-promoting point estimates, we identified five exploratory mediation patterns with positive indirect-effect estimates along the putative gut microbiota-IID-cardiovascular disease pathway (Supplementary Data 20 and 21; Supplementary Fig. S11).

In the German microbiome GWAS, which comprised 430 microbial features in 8,956 participants, genetically predicted abundance of *OTU99_123*, classified as *Phascolarctobacterium*, showed a positive indirect association with atrial fibrillation and flutter through IID. The estimated mediation proportion was 9.05% (Supplementary Data 21; Supplementary Fig. S11d).

In the Dutch Microbiome Project (DMP) microbiome dataset, which included 412 microbial features, *Lachnospiraceae bacterium* 5_1_63FAA showed a positive indirect association with myocardial infarction through IID, with an estimated mediation proportion of 7.26% (Supplementary Data 21; Supplementary Fig. S11e). The MiBioGen consortium dataset included 24 cohorts, 18,340 participants, and 211 microbial taxonomic features. In this dataset, *Oxalobacter* was implicated in three positive mediation pathways: *Oxalobacter*-IID-cardiac arrhythmias, with a mediation proportion of 10.32%; *Oxalobacter*-IID-myocardial infarction, with a mediation proportion of 6.58%; and *Oxalobacter*-IID-atrial fibrillation and flutter, with a mediation proportion of 8.18% (Supplementary Data 21; Supplementary Fig. S11a-c).

Across all five pathways, the microbial trait was positively associated with IID, and IID was positively associated with the corresponding cardiovascular outcome, resulting in positive indirect effects. Higher genetically predicted abundance of *Phascolarctobacterium* was associated with increased liability to IID (β = 0.0337), which was subsequently associated with a higher risk of atrial fibrillation and flutter (β = 0.1027). Similarly, *Lachnospiraceae bacterium 5_1_63FAA* was positively associated with IID (β = 0.0379), whereas IID was positively associated with myocardial infarction (β = 0.0924). In the MiBioGen dataset, *Oxalobacter* showed a consistent positive association with IID (β = 0.0496), producing positive indirect effects for all three cardiovascular outcomes (Supplementary Data 21; Supplementary Fig. S11).

Among the five positive exploratory mediation patterns, only *Oxalobacter*-IID-cardiac arrhythmias reached nominal significance for the indirect effect (0.00542; 95% CI 0.00023 to 0.01062; two-sided, *P* = 0.0407). The remaining four indirect-effect estimates did not reach unadjusted *P* < 0.05: OTU99_123 (*Phascolarctobacterium*) abundance-IID-atrial fibrillation and flutter (0.00347; 95% CI −0.00054 to 0.00747; *P* = 0.0899); *Lachnospiraceae bacterium* 5_1_63FAA-IID-myocardial infarction (0.00350; 95% CI −0.00072 to 0.00772; *P* = 0.1038); *Oxalobacter*-IID-myocardial infarction (0.00459; 95% CI −0.00089 to 0.01007; *P* = 0.1010); and *Oxalobacter*-IID-atrial fibrillation and flutter (0.00510; 95% CI −0.00092 to 0.01112; *P* = 0.0967) (Supplementary Data 21).

*Oxalobacter* recurred across cardiac arrhythmias, myocardial infarction, and atrial fibrillation and flutter among the positive mediation patterns. These three outcome-specific analyses shared the same upstream *Oxalobacter*-to-IID estimate, and their recurrence therefore reflects a common upstream component rather than independent replication. Taken together, the analyses highlighted putative gut microbiota-IID-cardiovascular disease mediation patterns, although statistical uncertainty limits conclusions about the presence and magnitude of mediation (Supplementary Fig. S11). Protective concordant patterns and directionally inconsistent patterns are listed in the Pattern column of Supplementary Data 21.

## Discussion

In this study, we systematically investigated the shared genetic architecture between digestive infectious diseases and cardiovascular diseases using an integrated summary-statistics-based framework. After excluding seven traits with negative or near-zero observed-scale heritability estimates, 16 of the initial 36 traits were retained for downstream analyses, including three digestive infectious disease traits and 13 cardiovascular traits. All 39 prioritised digestive infection-cardiovascular disease pairs showed significant global genetic correlations after Bonferroni correction. At the local level, SUPERGNOVA identified 5 to 11 regions with *P* < 0.05 across IID-cardiovascular disease pairs; however, these signals remained exploratory because none survived block-level multiple-testing correction. MTAG identified 41 trait-pair-specific pleiotropic SNP records and 14 trait-pair-specific independent lead-SNP records, including 39 records and 12 lead variants involving IID.

Fine-mapping and colocalisation yielded eight candidate trait-pair-variant records representing four unique variants: rs486632, rs579459, rs653178 and rs17630235. These records showed concordant support across PLACO+, CPMA, CPASSOC-SHet and GPA. Gene-level and functional analyses implicated cardiac electrophysiology, coagulation, thrombosis and ischaemic vascular biology. Bidirectional MR supported selective directional relationships between genetically proxied IID liability and cardiovascular outcomes, and the mediation analyses identified five putative gut microbiota-IID-cardiovascular disease pathways across three microbiome GWAS resources.

Taken together, these findings support shared inherited susceptibility and provide a set of candidate loci, genes and host-microbe pathways for further investigation. They should not be interpreted as evidence that an acute episode of intestinal infection directly causes cardiovascular disease in every individual.

### Widespread but structured genetic sharing

The observation that all 39 prioritised trait pairs showed significant global genetic correlations suggests that the genetic relationship between digestive infectious diseases and cardiovascular diseases is not restricted to a single cardiovascular endpoint. This finding is compatible with epidemiological evidence linking severe infections with subsequent cardiovascular events, although genetic liability to a disease phenotype is not equivalent to the short-term biological effects of an acute infection [6, 69]. Cross-trait LDSC estimates genome-wide covariance in genetic effects rather than clinical causality, and therefore the observed correlations may reflect shared biological pathways, correlated disease definitions or other features of the source GWAS datasets. A recent multivariate GWAS of autoimmune and cardiovascular diseases identified shared genetic signals enriched in arterial tissues and lipid metabolism pathways [70]. In the present study, IID-centred analyses prioritised candidate signals related to cardiac electrophysiology, coagulation and thrombosis.

The predominance of positive genetic correlations is consistent with partially shared mechanisms involving vascular injury, cardiac electrophysiology, coagulation and thrombosis. Nevertheless, the three negative correlations involving cerebrovascular disease highlight the importance of retaining the direction of the genetic correlation estimates. A negative genetic correlation should not automatically be interpreted as a protective biological relationship. It may reflect differences in phenotype definition, disease ascertainment, sample composition, medication use or the underlying genetic architecture. The cerebrovascular disease findings should therefore be regarded as hypothesis-generating and require independent replication. Large GWAS have identified susceptibility loci for stroke and its subtypes across ancestries, providing a basis for further evaluation of our cerebrovascular findings by disease subtype [71].

At the local level, SUPERGNOVA identified five to 11 regions with *P*<0.05 across the IID-related cardiovascular trait pairs. The number and genomic distribution of these regions varied between cardiovascular outcomes, and several cytoband intervals recurred across trait pairs. However, no local signal survived the predefined Bonferroni correction across the 1,703 genomic blocks. The observed pattern is therefore compatible with heterogeneous and potentially multiregional local genetic sharing, but does not establish definitive local pleiotropy. The recurrent regions may help prioritise future analyses, but should not be interpreted as confirmed local causal regions.

### IID as the central digestive infectious disease phenotype

The concentration of variant-level signals in IID-related pairs was a central feature of the study. Although three digestive infectious disease traits passed the univariate heritability criteria, IID accounted for 39 of the 41 pleiotropic SNP records and 12 of the 14 independent lead variants. This pattern supports the use of IID as the principal digestive infectious disease phenotype for subsequent fine-mapping, functional prioritisation and MR analyses, while also indicating heterogeneity across digestive infectious disease phenotypes.

Several explanations may account for this concentration. IID may represent a broader or more genetically heterogeneous phenotype than pathogen-specific diagnoses and may therefore capture a wider range of host susceptibility mechanisms. The IID GWAS may also have had greater statistical power or a different case-ascertainment structure. Differences in disease severity, diagnostic coding and sample size may further influence variant discovery. Accordingly, the predominance of IID should not be interpreted as evidence that the other digestive infectious disease traits are biologically irrelevant.

MTAG can improve power by jointly modelling genetic associations across traits, but its performance depends on assumptions concerning cross-trait effect patterns and sample-statistics properties. The more favourable MTAG maxFDR estimates observed for IID-related pairs provide supportive calibration evidence, but they do not constitute an independent estimate of the false discovery rate. The IID-centred downstream strategy should therefore be viewed as a biologically and statistically motivated prioritisation, rather than as evidence that the same mechanism applies uniformly to all digestive infections.

### Candidate loci and biological interpretation

The recurrent loci identified in this study mapped to biologically plausible cardiovascular pathways. The 4q25 region contained *PITX2* and *ENPEP* signals in arrhythmia-related trait pairs. The 4q25 region and *PITX2* have been repeatedly implicated in atrial fibrillation and atrial electrophysiology [72–74]. *ENPEP* has also been discussed in relation to angiotensin metabolism and vascular biology [75]. In rats, inhibition of central aminopeptidase A reduced angiotensin III formation and lowered blood pressure [76]. Thus, the recurrence of this region in the IID–cardiac arrhythmia and IID–AF pairs is consistent with a shared electrophysiological component. However, the present studies haven’t demonstrated that *PITX2* or *ENPEP* is the causal mediator of the association.

The 9q34.2 region was particularly prominent in the IID-venous thromboembolism analysis. The prioritised genes included *ABO* and *ADAMTS13*, which are biologically relevant to coagulation, platelet biology and thrombotic risk. Pathogenic *ADAMTS13* variants cause thrombotic thrombocytopenic purpura, demonstrating the importance of von Willebrand factor processing in thrombotic biology [77]. The *ABO* PWAS association provided additional protein-level support for this region, and previous genetic studies have linked the *ABO* region to venous thromboembolism risk [78, 79]. Nevertheless, the combination of proximity, MAGMA association, TWAS and PWAS evidence does not uniquely identify a causal gene. These results should therefore be interpreted as convergent gene-prioritisation evidence.

The 12q24.12-q24.13 region contained signals involving *SH2B3*, *ATXN2*, *NAA25*, *ALDH2*, *BRAP* and *ACAD10*. The region has previously been implicated in coronary artery disease and cardiometabolic biology, including signals involving *BRAP* and *ALDH2* [80]. Its recurrence in the IID-ischaemic heart disease analysis is compatible with a possible immune-vascular or cardiometabolic component, but the present study did not directly establish the molecular pathway linking IID to this region.

Large-scale coronary artery disease GWAS have expanded the catalogue of susceptibility loci and candidate genes, providing an external context for the coronary-related signals prioritised here [81].

The functional analyses indicated outcome-specific biological patterns. Gene-level annotations and tissue-specific enrichment implicated cardiac electrophysiology, whereas coagulation, thrombosis and lipid-related processes were prominent in the vascular analyses. GSEA identified 43 significant trait-pair-GO term associations across IID-venous thromboembolism, IID-myocardial infarction and IID-ischaemic heart disease; no KEGG pathway met the prespecified criteria (Supplementary Data 11). TSEA identified seven significant tissue-pair associations after within-pair correction across 47 tissues (Supplementary Data 12). The adult entries with combined *P* < 0.001 were nominally significant (Supplementary Data 13). TWAS identified 21 candidate genes with at least one FDR-significant association, while PWAS identified four associations involving *ABO*, *OBP2B* and *ENPEP* (Supplementary Data 15 and 16). These results provide complementary gene-prioritisation evidence, but genetically predicted expression or protein associations do not establish tissue-specific mediation or gene-level causality [82, 83]. Large plasma proteomic resources, such as the UK Biobank Pharma Proteomics Project, provide opportunities to evaluate these protein-level candidates in additional datasets [84].

### Evidence for potential directional relationships from bidirectional MR

The MR findings varied across cardiovascular outcomes and directions of analysis, with only the IID–cardiac arrhythmias pair showing associations that met the FDR-adjusted significance threshold in both directions.Genetically proxied IID liability was associated with cardiac arrhythmias, myocardial infarction, atrial fibrillation and flutter, and ischaemic heart disease, whereas the reverse analysis supported an association only between cardiac arrhythmia liability and IID. IID and cardiac arrhythmias therefore represented the only pair with statistically supported associations in both directions (Supplementary Data 18).

These findings should not be interpreted as definitive evidence that IID directly causes cardiovascular disease. MR estimates represent the effect of genetically proxied liability under instrumental-variable assumptions and may not correspond to the effect of an acute clinical infection [85]. Horizontal pleiotropy remains possible, particularly because variants associated with IID may also influence immune, vascular or metabolic traits through pathways independent of the analysed cardiovascular outcomes. Invalid instruments can bias MR estimates even when conventional sensitivity analyses do not identify a clear violation [58, 86]. Potential participant overlap between exposure and outcome GWAS may further bias MR estimates, particularly when instruments are weak [87].

The attenuation of the IID-myocardial infarction association after exclusion of variants associated with potential confounding phenotypes illustrates the importance of sensitivity analyses. Conversely, the persistence of the IID-arrhythmia and IID-ischaemic heart disease associations after several exclusions increases confidence in their directional robustness, but does not eliminate all forms of pleiotropic bias (Supplementary Data 18). The results should therefore be reported as evidence supporting potential directional relationships rather than definitive clinical causation, in keeping with current MR reporting recommendations [88].

### Putative gut microbiota-mediated pathways

The mediation analyses identified five positive gut microbiota-IID-cardiovascular disease pathways across three microbiome GWAS resources. The recurrence of *Oxalobacter* across cardiac arrhythmias, myocardial infarction and atrial fibrillation and flutter was particularly notable (Supplementary Data 21). Previous MR work has also reported an association between genetically predicted *Oxalobacter* and coronary artery disease, providing relevant biological context for the present findings. However, the earlier and current analyses are not independent replication studies of the same endpoint and should not be interpreted as definitive confirmation.

The microbiome findings are biologically compatible with several possible mechanisms, including intestinal barrier function, immune signalling, host metabolism and microbial metabolites. Previous experimental and clinical studies have linked gut microbial metabolism, including trimethylamine N-oxide-related pathways, with cardiovascular risk [23, 24, 89]. Experimental work has shown that TMAO can increase platelet reactivity and thrombosis potential [90]. These mechanisms provide context for the present results but were not directly measured in this study. Therefore, the current findings do not establish that the identified microbial taxa act through a specific metabolite or inflammatory pathway.

The mediation proportions should also be interpreted cautiously. They were derived from summary-level genetic associations and depend on the assumptions of two-step MR mediation analysis. Microbiome GWAS datasets differ in sequencing platforms, taxonomic resolution, cohort composition and phenotype processing, and host-microbiome associations can vary substantially across cohorts [17, 91]. The use of suggestive instrument thresholds to improve coverage may increase weak-instrument and horizontal-pleiotropy bias. In addition, mediated proportions may be unstable when the total effect is small or when direct and indirect effects have different directions. The five pathways should therefore be regarded as putative genetically mediated pathways requiring independent longitudinal and experimental validation.

### Strengths and limitations

This study has several strengths. We evaluated a broad set of digestive infectious and cardiovascular phenotypes using a staged analytical framework. We integrated univariate and cross-trait LDSC, local genetic covariance analysis, MTAG, complementary pleiotropy methods, fine-mapping, colocalisation, MAGMA, tissue- and cell-type-specific enrichment, TWAS, PWAS, bidirectional MR and microbiome mediation analysis. The distinction between trait-pair-specific records and unique variants prevented repeated signals across multiple trait pairs from being incorrectly counted as independent variants. In addition, the use of three microbiome GWAS resources allowed the consistency of putative mediation patterns to be assessed across different datasets.

Several limitations should be acknowledged. First, all analyses were based on GWAS summary statistics, which prevented individual-level assessment of age, sex, disease severity, medication use, comorbidity, infection timing and other clinical characteristics. Heterogeneity in phenotype definition, case ascertainment, prevalence and source-study design may therefore have influenced the results.

Second, most GWAS datasets were derived from European-ancestry populations, limiting generalisability to other ancestries.

Third, the exclusion of seven traits with negative or near-zero observed-scale heritability estimates and the subsequent IID-centred prioritisation may have reduced the generalisability of the downstream findings to pathogen-specific digestive infections. The trait-level filter was intended to improve statistical stability, but it may also have preferentially retained traits with stronger GWAS signals.

Fourth, MTAG [14], fine-mapping and colocalisation are model-dependent. Results may be affected by incomplete variant coverage, the quality of the LD reference panel, allelic heterogeneity and the assumption of at most one causal signal in the coloc model [15, 41]. Fifth, the ±500-kb gene-annotation window may miss distal regulatory relationships, while MAGMA, TWAS and PWAS associations may reflect linkage disequilibrium or correlated regulatory effects rather than direct gene-level causality. Finally, MR and mediation analyses remain subject to weak-instrument bias, horizontal pleiotropy, winner’s curse and measurement heterogeneity. Cross-method sensitivity analyses provide methodological concordance but do not replace independent replication in external cohorts. The microbiome mediation results were additionally affected by taxonomic heterogeneity and the use of a genome-wide suggestive instrument threshold. Experimental studies, larger multi-ancestry GWAS, longitudinal infection cohorts, metagenomic and metabolomic profiling, and independent clinical replication will be required to establish the biological and clinical relevance of the identified pathways.

### Implications

The findings provide a genetic prioritisation framework for studying how intestinal infectious disease liability may intersect with cardiovascular disease biology. The recurrent signals at electrophysiological, coagulation, thrombosis and immune-vascular loci identify candidate regions for functional investigation. The microbiome findings further suggest that host-microbe interactions may contribute to selected IID-cardiovascular disease relationships.

These results should not be interpreted as an immediate basis for microbiome intervention, drug repurposing or clinical risk prediction. In particular, the druggability annotations for *ENPE*P and *OBP2B* identify therapeutic hypotheses rather than validated treatment targets. Future studies should combine multi-ancestry genetic data with longitudinal infection phenotypes, metagenomic and metabolomic measurements, cell-based experiments, genome editing and prospective clinical validation.

## Conclusions

In conclusion, digestive infectious diseases and cardiovascular diseases showed widespread, directionally structured genetic sharing in this European-ancestry analysis. IID emerged as the principal digestive infectious disease phenotype contributing to the cross-trait pleiotropic signals. Integrative analyses prioritised recurrent candidate variants and genes related to cardiac electrophysiology, coagulation, thrombosis and ischaemic vascular biology. Bidirectional Mendelian randomisation provided evidence consistent with potential directional associations between IID liability and selected cardiovascular outcomes; evidence in both directions was observed only for IID and cardiac arrhythmias. Mediation analyses identified five putative gut microbiota-IID-cardiovascular disease pathways across three microbiome GWAS resources. These findings provide a genetic perspective complementary to epidemiological evidence linking acute infections with cardiovascular events, and to previous genetic and mechanistic studies of gut microbial variation and cardiovascular biology. The prioritised loci, genes and microbial pathways offer testable hypotheses for future functional and clinical studies.

## List of abbreviations

AET: Arterial embolism and thrombosis
AF: Atrial fibrillation and flutter
ARIC: Atherosclerosis Risk in Communities
CAD: Coronary artery disease
CPASSOC: Cross-phenotype association
CPMA: Cross-phenotype meta-analysis
CSEA: Cell-type-specific enrichment analysis
CVD: Cardiovascular disease
EUR: European ancestry
FDR: False discovery rate
GO: Gene Ontology
GPA: Genetic analysis incorporating Pleiotropy and Annotation
GSEA: Gene-set enrichment analysis
GTEx: Genotype-Tissue Expression
GWAS: Genome-wide association study
HF: Heart failure
IID: Intestinal infectious diseases
IHD: Ischaemic heart disease
IV: Instrumental variable
IVW: Inverse-variance-weighted
KEGG: Kyoto Encyclopedia of Genes and Genomes
LD: Linkage disequilibrium
LDSC: Linkage disequilibrium score regression
MAGMA: Multi-marker analysis of genomic annotation
MHC: Major histocompatibility complex
MI: Myocardial infarction
MR: Mendelian randomisation
MR-PRESSO: Mendelian randomisation pleiotropy residual sum and outlier
MR-RAPS: Mendelian randomisation robust adjusted profile score
MTAG: Multi-trait analysis of GWAS
NES: Normalised enrichment score
OR: Odds ratio
PE: Pulmonary embolism
PLACO+: Pleiotropic analysis under a composite null hypothesis with adjustment for cross-trait correlation
PPH4: Posterior probability of hypothesis H4 (a shared causal variant for both traits)
pQTL: Protein quantitative trait locus
PWAS: Proteome-wide association study
RVD: Rheumatic valve diseases
SE: Standard error
SNP: Single-nucleotide polymorphism
TSEA: Tissue-specific enrichment analysis
TTD: Therapeutic Target Database
TWAS: Transcriptome-wide association study
VTE: Venous thromboembolism
(h2): SNP heritability
(rg): Genetic correlation
(Z): Heritability Z score (used for quality control in Fig. 1)

## Declarations

### Ethics approval and consent to participate

Ethics declaration: not applicable. This study involved secondary analyses of publicly available GWAS summary statistics. No additional ethics approval was required under the applicable institutional policy.

Consent to Participate declaration: not applicable. No participants were recruited and no new participant data were collected for this study.

### Consent for publication

Consent to Publish declaration: not applicable. The manuscript does not contain any identifiable participant information.

### Availability of data and materials

All GWAS summary statistics analysed in this study were obtained from publicly available resources. The disease GWAS summary statistics used in the cross-trait analyses were obtained from the disease-centric Mendelian randomization database (DMRdb). GWAS identifiers, source studies, sample sizes and data-access information for the disease traits are provided in Supplementary Data 1. The characteristics of the FinnGen Release 10 datasets used in the bidirectional Mendelian randomisation analyses are summarised in Supplementary Data 17.

Source publications for the German, Dutch Microbiome Project (DMP) and MiBioGen gut microbiome GWAS are cited in the Methods. Public reference datasets and prediction models used in the analyses included the HapMap3 SNP set, European-ancestry LD-score files, 1000 Genomes Project Phase 3 European-ancestry reference genotypes, LDetect/Fourier genomic partitions, GTEx v8/FUSION expression weights and ARIC plasma-protein models; the corresponding resources are identified in the Methods and associated references. Study-generated results are provided in the accompanying Supplementary Data files. No individual-level phenotype or genotype data from the source disease or microbiome GWAS participants were accessed. Public 1000 Genomes Project Phase 3 genotype data were used solely as a reference panel for LD-based analyses.

## Authors’ contributions

JN, YC and JP conceived and designed the study. JN collected and curated the data, performed the statistical and bioinformatics analyses, and drafted the manuscript. YC and JP supervised the study and critically revised the manuscript. All authors reviewed and approved the final manuscript.

## Supporting information

Supplemental Data 21, and will be used for the link to the file on the preprint site.

Supplemental Data 01, and will be used for the link to the file on the preprint site.

Supplemental Data 02, and will be used for the link to the file on the preprint site.

Supplemental Data 03, and will be used for the link to the file on the preprint site.

Supplemental Data 04, and will be used for the link to the file on the preprint site.

Supplemental Data 05, and will be used for the link to the file on the preprint site.

Supplemental Data 06, and will be used for the link to the file on the preprint site.

Supplemental Data 07, and will be used for the link to the file on the preprint site.

Supplemental Data 08, and will be used for the link to the file on the preprint site.

Supplemental Data 09, and will be used for the link to the file on the preprint site.

Supplemental Data 10, and will be used for the link to the file on the preprint site.

Supplemental Data 11, and will be used for the link to the file on the preprint site.

Supplemental Data 12, and will be used for the link to the file on the preprint site.

Supplemental Data 13, and will be used for the link to the file on the preprint site.

Supplemental Data 14, and will be used for the link to the file on the preprint site.

Supplemental Data 15, and will be used for the link to the file on the preprint site.

Supplemental Data 16, and will be used for the link to the file on the preprint site.

Supplemental Data 17, and will be used for the link to the file on the preprint site.

Supplemental Data 18, and will be used for the link to the file on the preprint site.

Supplemental Data 19, and will be used for the link to the file on the preprint site.

Supplemental Data 20, and will be used for the link to the file on the preprint site.

Supplemental Fig S1, and will be used for the link to the file on the preprint site.

Supplemental Fig S2, and will be used for the link to the file on the preprint site.

Supplemental Fig S3, and will be used for the link to the file on the preprint site.

Supplemental Fig S4, and will be used for the link to the file on the preprint site.

Supplemental Fig S5, and will be used for the link to the file on the preprint site.

Supplemental Fig S6, and will be used for the link to the file on the preprint site.

Supplemental Fig S7, and will be used for the link to the file on the preprint site.

Supplemental Fig S8, and will be used for the link to the file on the preprint site.

Supplemental Fig S9, and will be used for the link to the file on the preprint site.

Supplemental Fig S10, and will be used for the link to the file on the preprint site.

Supplemental Fig S11, and will be used for the link to the file on the preprint site.

## Data Availability

All GWAS summary statistics analysed in this study were obtained from publicly available resources. The disease GWAS summary statistics used in the cross-trait analyses were obtained from the disease-centric Mendelian randomization database (DMRdb). GWAS identifiers, source studies, sample sizes and data-access information for the disease traits are provided in Supplementary Data 1. The characteristics of the FinnGen Release 10 datasets used in the bidirectional Mendelian randomisation analyses are summarised in Supplementary Data 17. Source publications for the German, Dutch Microbiome Project (DMP) and MiBioGen gut microbiome GWAS are cited in the Methods. Public reference datasets and prediction models used in the analyses included the HapMap3 SNP set, European-ancestry LD-score files, 1000 Genomes Project Phase 3 European-ancestry reference genotypes, LDetect/Fourier genomic partitions, GTEx v8/FUSION expression weights and ARIC plasma-protein models; the corresponding resources are identified in the Methods and associated references. Study-generated results are provided in the accompanying Supplementary Data files. No individual-level phenotype or genotype data from the source disease or microbiome GWAS participants were accessed. Public 1000 Genomes Project Phase 3 genotype data were used solely as a reference panel for LD-based analyses.

## Acknowledgements

We thank the participants and investigators of the original disease and gut microbiome genome-wide association studies for making their summary statistics available. We acknowledge the participants and investigators of the FinnGen study, the German and Dutch Microbiome Project (DMP) microbiome GWAS teams, the MiBioGen consortium, the GTEx project, the ARIC study and the 1000 Genomes Project for providing the data and reference resources used in this research. We also thank the GWAS Catalog team for curating the association data used to identify variants associated with potential confounding phenotypes, and the developers of the publicly available analytical software used in this study.

## Supplementary material

### Supplementary tables

**Supplementary Data 1.** Overview of the 16 digestive infectious traits and 20 cardiovascular traits in the EUR population included in this study.

**Supplementary Data 2.** Global genetic correlations between digestive infectious diseases and cardiovascular traits estimated using LDSC.

**Supplementary Data 3.** Nominal local genetic covariance regions between digestive infectious diseases and cardiovascular traits.

**Supplementary Data 4.** Candidate SNP records from cross-trait MTAG analysis. **Supplementary Data 5.** Trait-pair-level summary of cross-trait MTAG results and diagnostic metrics.

**Supplementary Data 6.** Independent pleiotropic genomic loci associated with the risk of digestive infectious diseases and cardiovascular diseases.

**Supplementary Data 7.** Fine-mapping and colocalisation results for independent pleiotropic loci.

**Supplementary Data 8.** Candidate pleiotropic variants supported by fine-mapping and colocalisation.

**Supplementary Data 9.** Cross-trait sensitivity analyses of candidate pleiotropic variants.

**Supplementary Data 10.** MAGMA gene-based associations for intestinal infectious disease–cardiovascular disease trait pairs.

**Supplementary Data 11.** Enriched GO terms identified by GSEA across the five prioritised IID–cardiovascular disease trait pairs.

**Supplementary Data 12.** Tissue-specific enrichment results across five prioritised IID–cardiovascular trait pairs.

**Supplementary Data 13.** Complete WebCSEA enrichment results across five prioritised IID–cardiovascular trait pairs.

**Supplementary Data 14.** Candidate pleiotropic genes prioritised by MAGMA within ±500 kb of candidate pleiotropic variants.

S**upplementary Data 15.** Significant TWAS results based on 24 candidate pleiotropic genes.

**Supplementary Data 16.** Significant PWAS results and druggability annotations for candidate pleiotropic genes.

**Supplementary Data 17.** Characteristics of GWAS datasets used in bidirectional Mendelian randomisation analyses.

**Supplementary Data 18.** Bidirectional Mendelian randomisation estimates across five IID–cardiovascular trait pairs.

**Supplementary Data 19.** Characteristics of genetic instruments used in bidirectional Mendelian randomisation analyses across five IID–cardiovascular trait pairs.

**Supplementary Data 20.** Mendelian randomisation estimates for gut microbial features in the IID–cardiovascular mediation analyses.

**Supplementary Data 21.** Two-step Mendelian randomisation estimates of gut microbiota–IID–cardiovascular mediation effects.

### Supplementary figures

**Supplementary Fig. S1.** Pair-specific pleiotropic lead-SNP records identified by MTAG.

**Supplementary Fig. S2.** Regional association plots for the eight final candidate trait-pair–variant records.

**Supplementary Fig. S3.** Relationships between final candidate loci and IID–cardiovascular trait pairs.

**Supplementary Fig. S4**. Cross-method association statistics for final candidate pleiotropic variants.

**Supplementary Fig. S5.** Method-specific support for final candidate pleiotropic variants.

**Supplementary Fig. S6.** MAGMA gene-based associations and final candidate gene mapping.

**Supplementary Fig. S7.** Tissue-specific enrichment across five prioritised IID–cardiovascular trait pairs.

**Supplementary Fig. S8.** Adult cell-type enrichment across organ systems and five trait pairs.

**Supplementary Fig. S9.** Significant TWAS associations for candidate pleiotropic genes.

**Supplementary Fig. S10.** Leave-one-out sensitivity analyses of bidirectional IID–cardiovascular associations.

**Supplementary Fig. S11.** Gut microbiota–IID–cardiovascular mediation patterns.

