## Supplemental Fig S1, and will be used for the link to the file on the preprint site. for "Shared genetic architecture between digestive infectious diseases and cardiovascular diseases: a genome-wide cross-trait analysis"

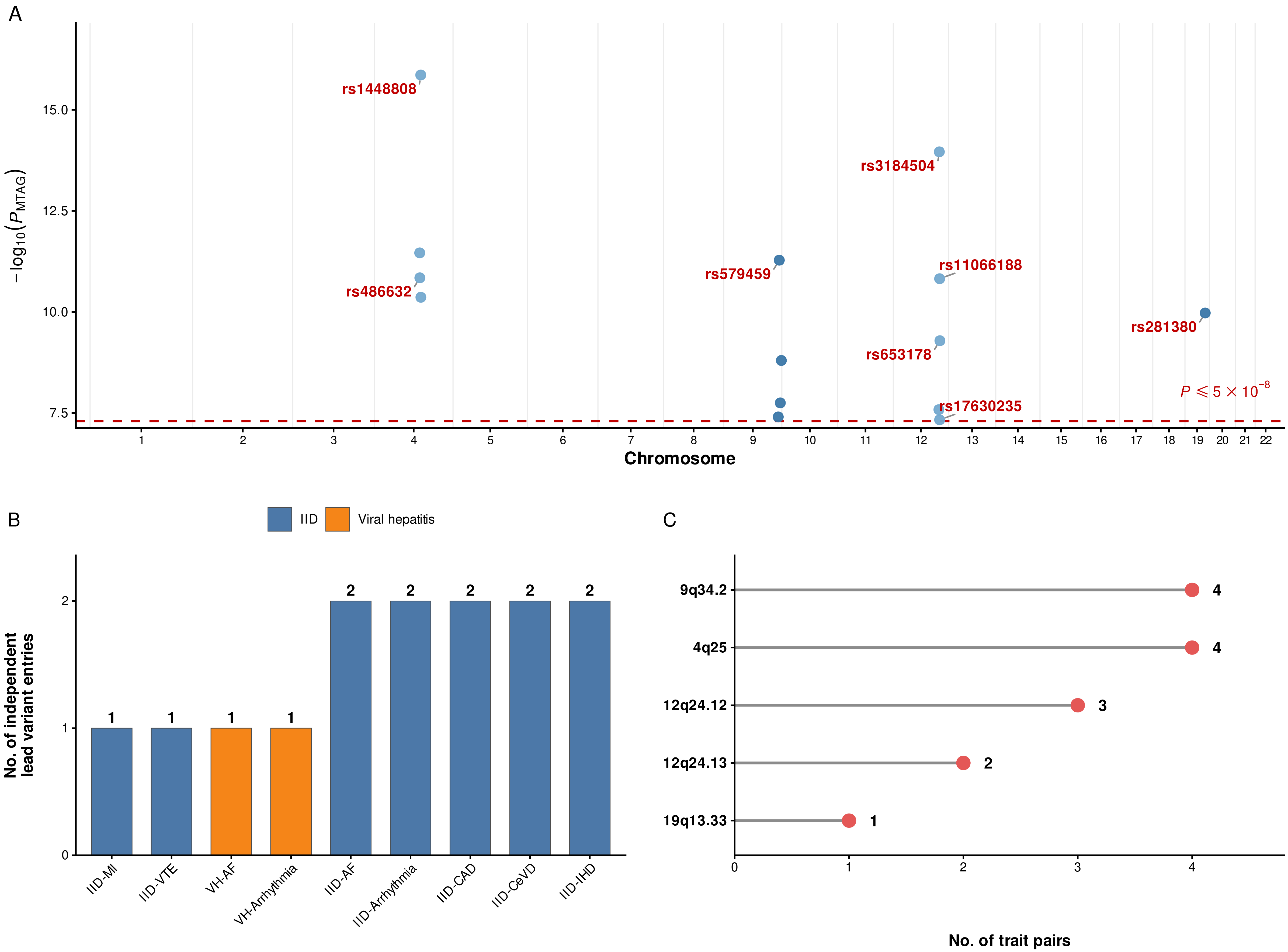

A
$-\log_{10}(P_{MTAG})$
15.0
12.5
10.0
7.5
rs1448808
rs3184504
rs486632
rs579459
rs11066188
rs281380
rs653178
rs17630235
$P \leqslant 5 \times 10^{-8}$
1 2 3 4 5 6 7 8 9 10 11 12 13 14 15 16 17 18 19 20 21 22
Chromosome
B
IID
Viral hepatitis
No. of independent lead variant entries
0
1
2
IID-MI
IID-VTE
VH-AF
VH-Arrhythmia
IID-AF
IID-Arrhythmia
IID-CAD
IID-CeVD
IID-IHD
C
9q34.2
4q25
12q24.12
12q24.13
19q13.33
0 1 2 3 4
No. of trait pairs
