## Supplemental Fig S2, and will be used for the link to the file on the preprint site. for "Shared genetic architecture between digestive infectious diseases and cardiovascular diseases: a genome-wide cross-trait analysis"

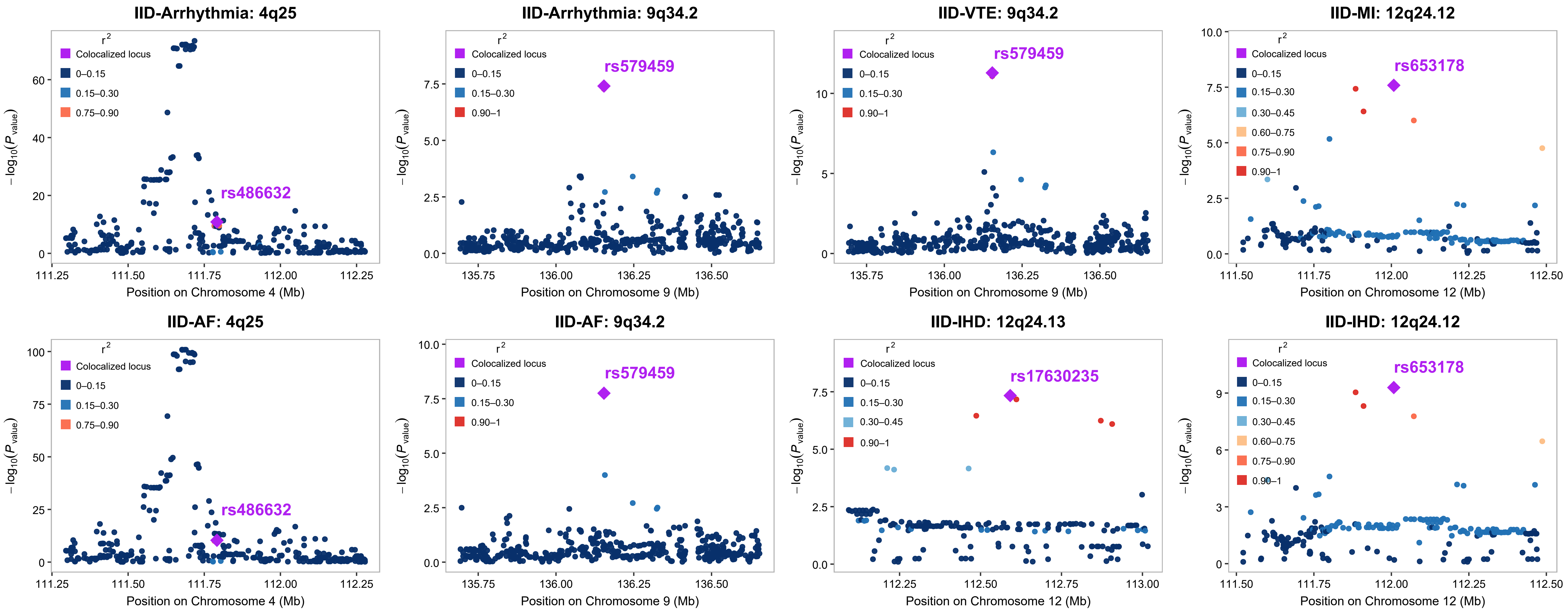

IID-Arrhythmia: 4q25
IID-Arrhythmia: 9q34.2
IID-VTE: 9q34.2
IID-MI: 12q24.12
IID-AF: 4q25
IID-AF: 9q34.2
IID-IHD: 12q24.13
IID-IHD: 12q24.12
r2
Colocalized locus
0–0.15
0.15–0.30
0.30–0.45
0.60–0.75
0.75–0.90
0.90–1
rs486632
rs579459
rs653178
rs17630235
−log10(P value)
Position on Chromosome 4 (Mb)
Position on Chromosome 9 (Mb)
Position on Chromosome 12 (Mb)
