## Supplementary figures and images for "Shared genetic architecture between digestive infectious diseases and cardiovascular diseases: a genome-wide cross-trait analysis"

### Supplemental Fig S3, and will be used for the link to the file on the preprint site.

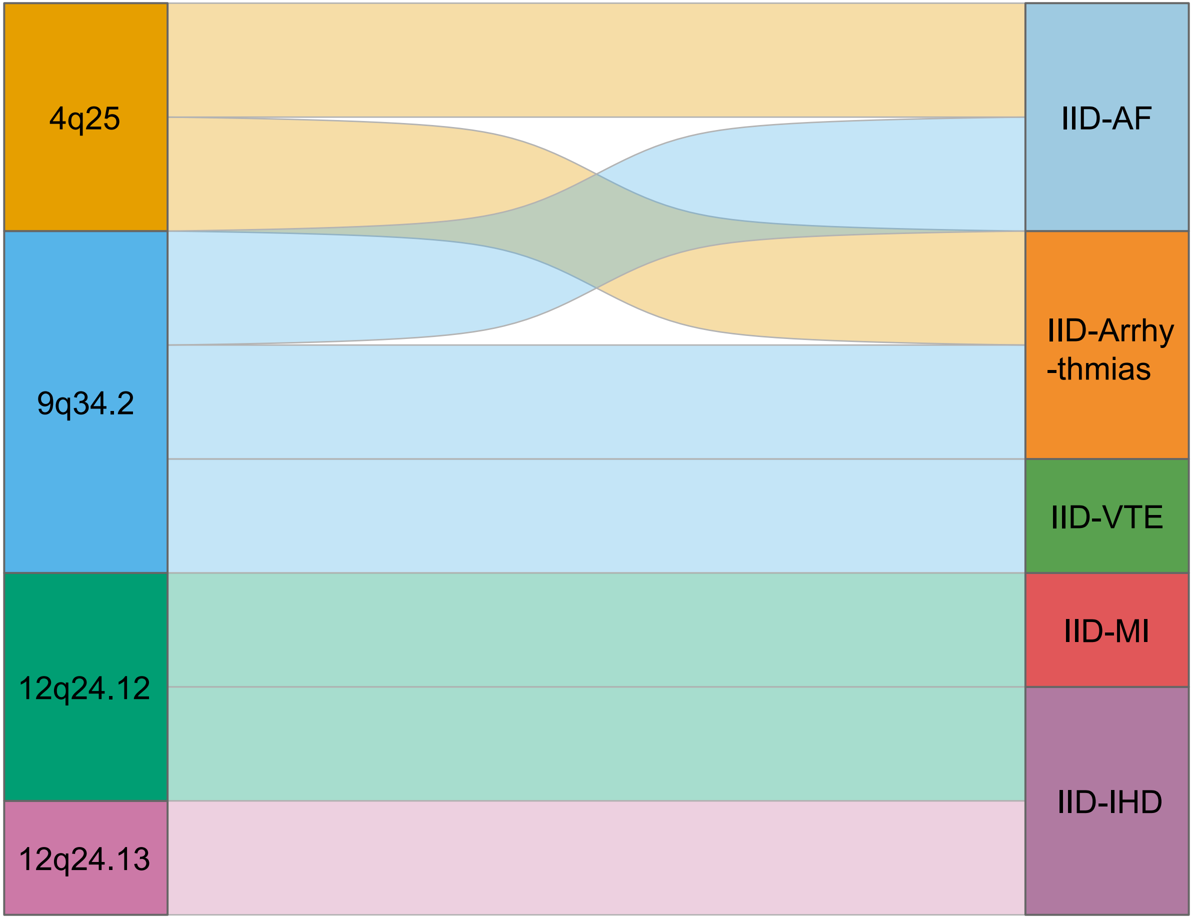

4q25
9q34.2
12q24.12
12q24.13
IID-AF
IID-Arrhy
-thmias
IID-VTE
IID-MI
IID-IHD
