## Supplemental Fig S4, and will be used for the link to the file on the preprint site. for "Shared genetic architecture between digestive infectious diseases and cardiovascular diseases: a genome-wide cross-trait analysis"

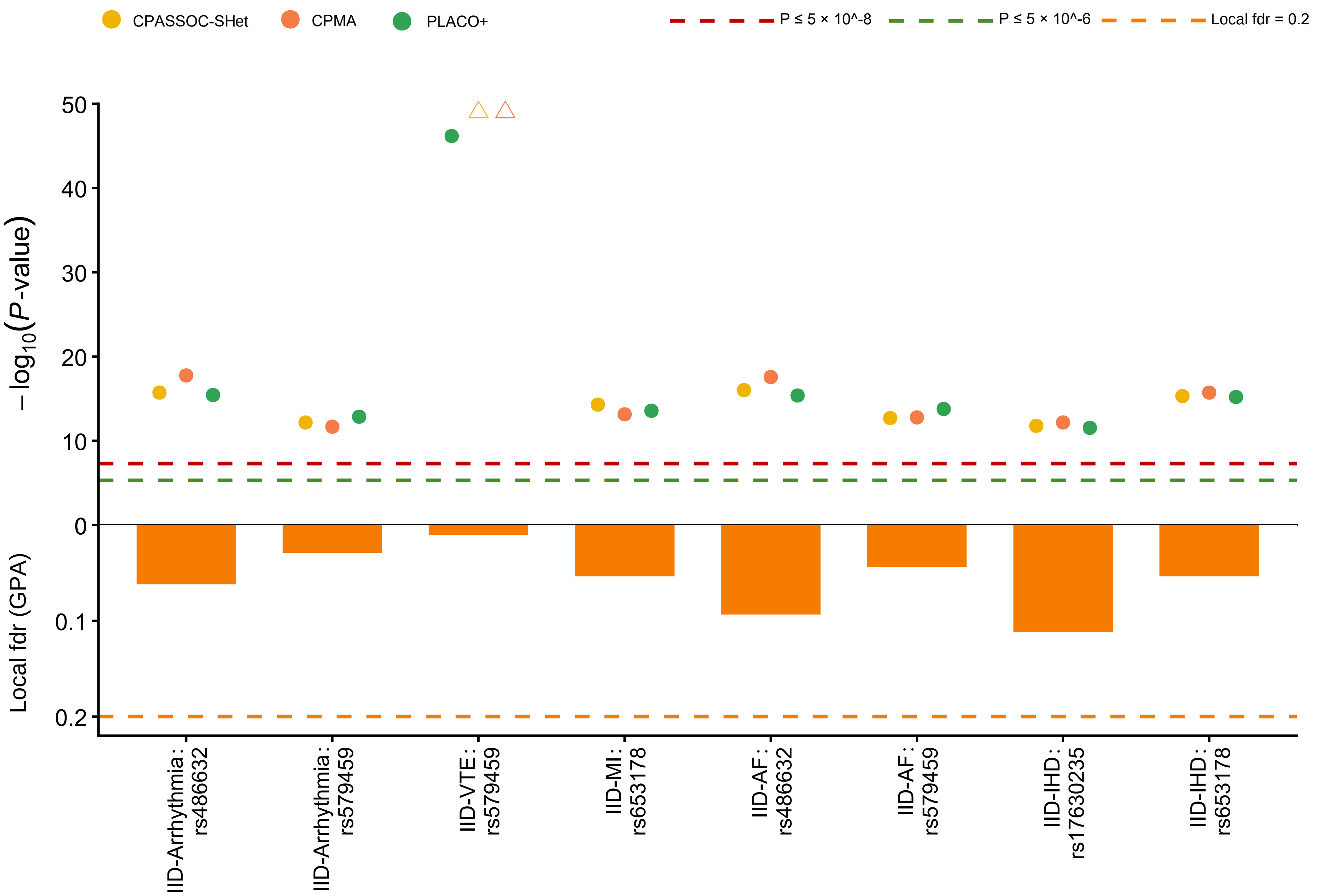

CPASSOC-SHet
CPMA
PLACO+
P ≤ 5 × 10^-8
P ≤ 5 × 10^-6
Local fdr = 0.2
$-\log_{10}(P\text{-value})$
Local fdr (GPA)
50
40
30
20
10
0
0.1
0.2
IID-Arrhythmia : rs486632
IID-Arrhythmia : rs579459
IID-VTE : rs579459
IID-MI : rs653178
IID-AF : rs486632
IID-AF : rs579459
IID-IHD : rs17630235
IID-IHD : rs653178
