## Supplemental Fig S5, and will be used for the link to the file on the preprint site. for "Shared genetic architecture between digestive infectious diseases and cardiovascular diseases: a genome-wide cross-trait analysis"

| Trait pairs | Candidate pleiotropic SNPs | Sample size | | Data source | |
|---|---|---|---|---|---|
| | | Intestinal infectious diseases | Cardiovascular diseases | Intestinal infectious diseases | Cardiovascular diseases |
| IID-Arrhythmia | rs486632 (4q25) | 412,181 | 313,778 | FinnGen | FinnGen |
| IID-Arrhythmia | rs579459 (9q34.2) | 412,181 | 313,778 | FinnGen | FinnGen |
| IID-VTE | rs579459 (9q34.2) | 412,181 | 412,181 | FinnGen | FinnGen |
| IID-MI | rs653178 (12q24.12) | 412,181 | 369,139 | FinnGen | FinnGen |
| IID-AF | rs486632 (4q25) | 412,181 | 261,395 | FinnGen | FinnGen |
| IID-AF | rs579459 (9q34.2) | 412,181 | 261,395 | FinnGen | FinnGen |
| IID-IHD | rs17630235 (12q24.13) | 412,181 | 412,181 | FinnGen | FinnGen |
| IID-IHD | rs653178 (12q24.12) | 412,181 | 412,181 | FinnGen | FinnGen |

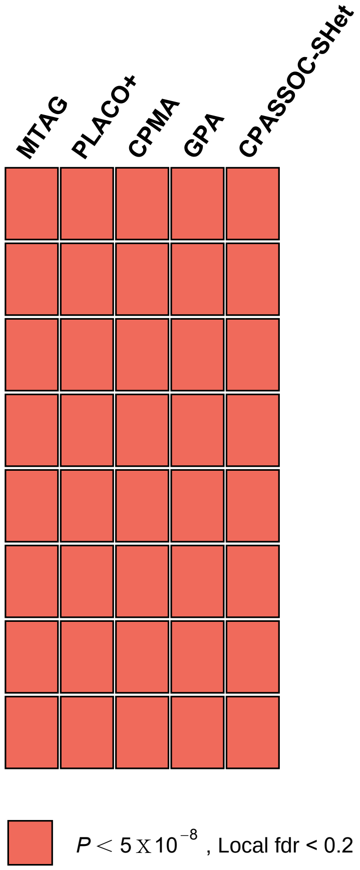
