## Supplemental Fig S6, and will be used for the link to the file on the preprint site. for "Shared genetic architecture between digestive infectious diseases and cardiovascular diseases: a genome-wide cross-trait analysis"

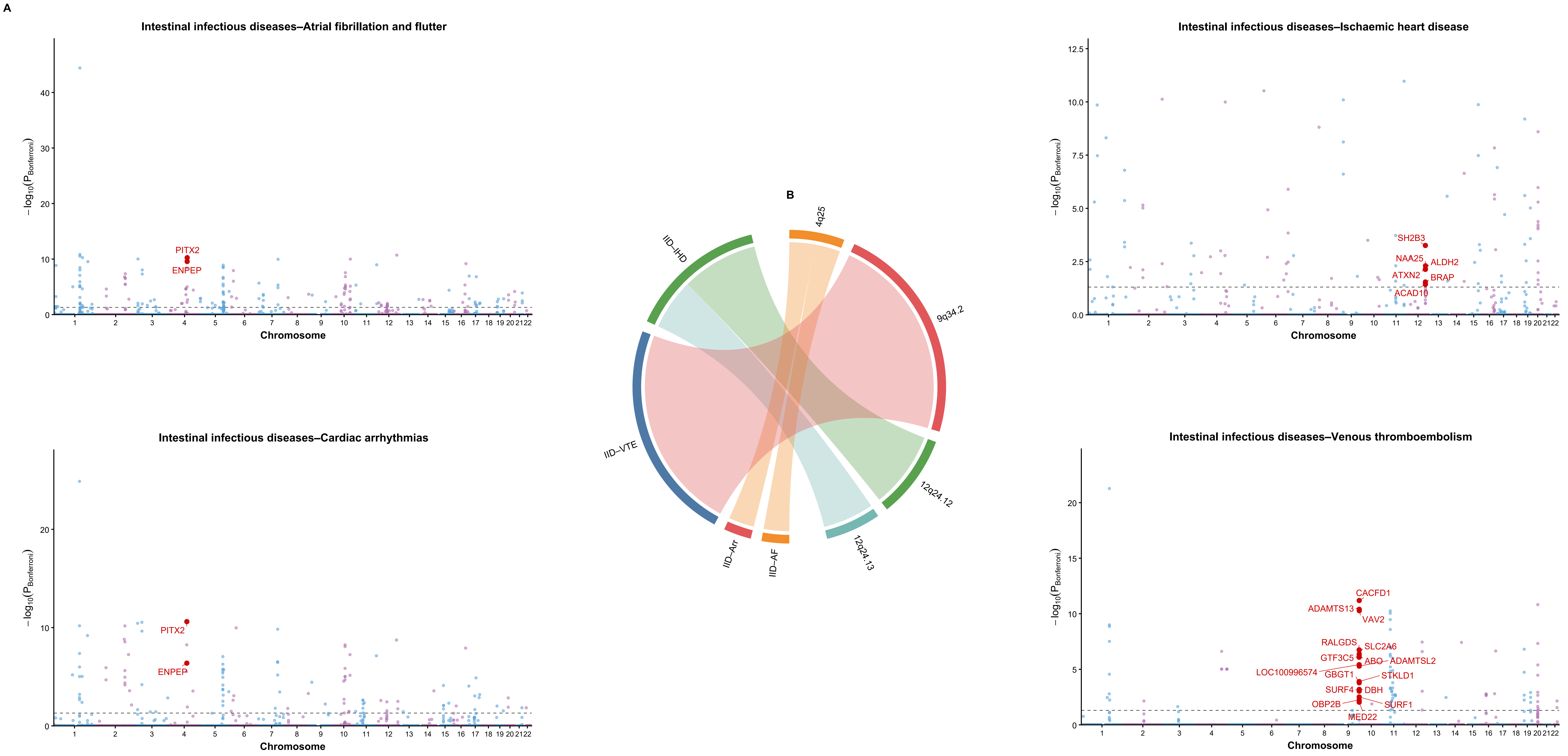

A
Intestinal infectious diseases–Atrial fibrillation and flutter
PITX2
ENPEP
Intestinal infectious diseases–Cardiac arrhythmias
PITX2
ENPEP
−log10(PBonferroni)
Chromosome
B
4q25
9q34.2
12q24.12
12q24.13
IID–AF
IID–Arr
IID–VTE
IID–IHD
Intestinal infectious diseases–Ischaemic heart disease
SH2B3
NAA25
ALDH2
ATXN2
BRAP
ACAD10
Intestinal infectious diseases–Venous thromboembolism
CACFD1
ADAMTS13
VAV2
RALGDS
SLC2A6
GTF3C5
ABO
ADAMTSL2
LOC100996574
GBGT1
STKLD1
SURF4
DBH
OBP2B
SURF1
MED22
