## Supplemental Fig S7, and will be used for the link to the file on the preprint site. for "Shared genetic architecture between digestive infectious diseases and cardiovascular diseases: a genome-wide cross-trait analysis"

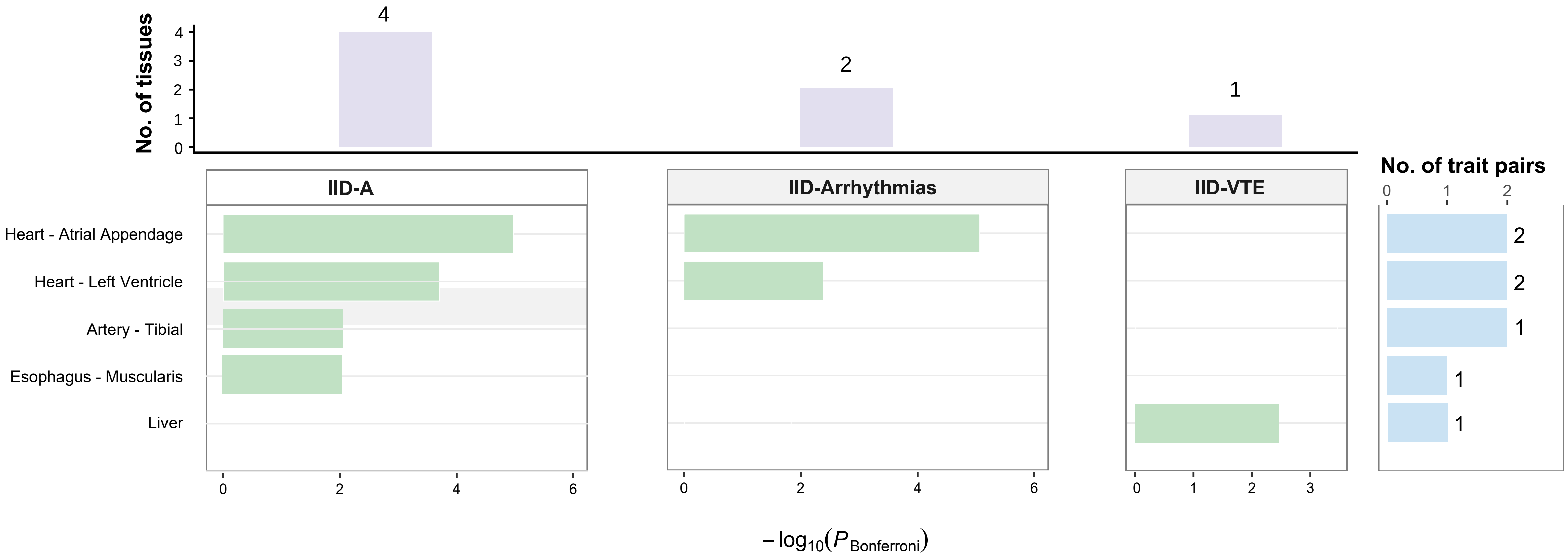
No. of tissues
4
2
1
IID-A
IID-Arrhythmias
IID-VTE
No. of trait pairs
Heart - Atrial Appendage
Heart - Left Ventricle
Artery - Tibial
Esophagus - Muscularis
Liver
2
2
1
1
1
$-\log_{10}(P_{\text{Bonferroni}})$
