## Supplemental Fig S8, and will be used for the link to the file on the preprint site. for "Shared genetic architecture between digestive infectious diseases and cardiovascular diseases: a genome-wide cross-trait analysis"

### Adult cell-type enrichment across five IID-cardiovascular trait pairs

4,705 entries; 941 adult reference panels per trait pair

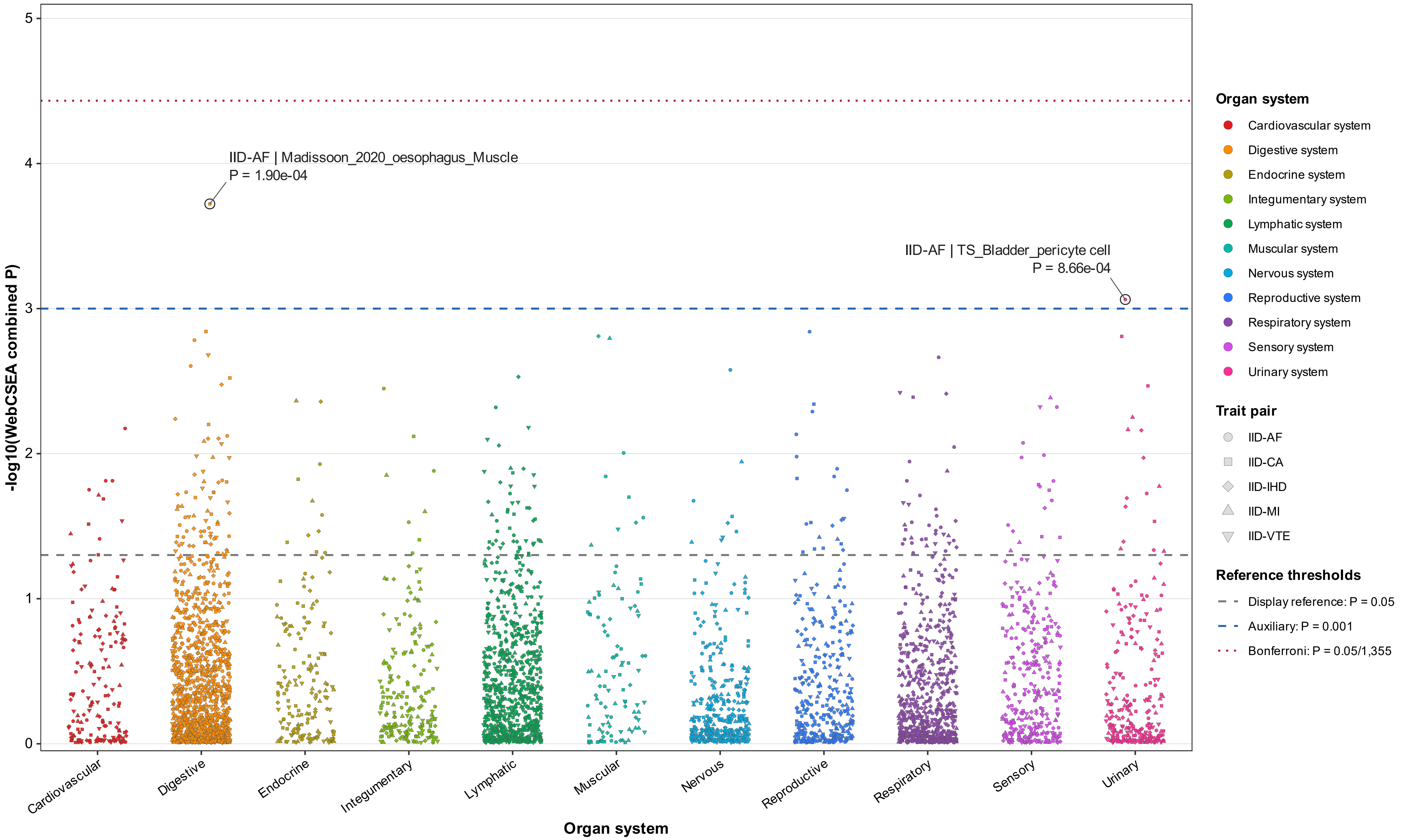
