## Supplemental Fig S9, and will be used for the link to the file on the preprint site. for "Shared genetic architecture between digestive infectious diseases and cardiovascular diseases: a genome-wide cross-trait analysis"

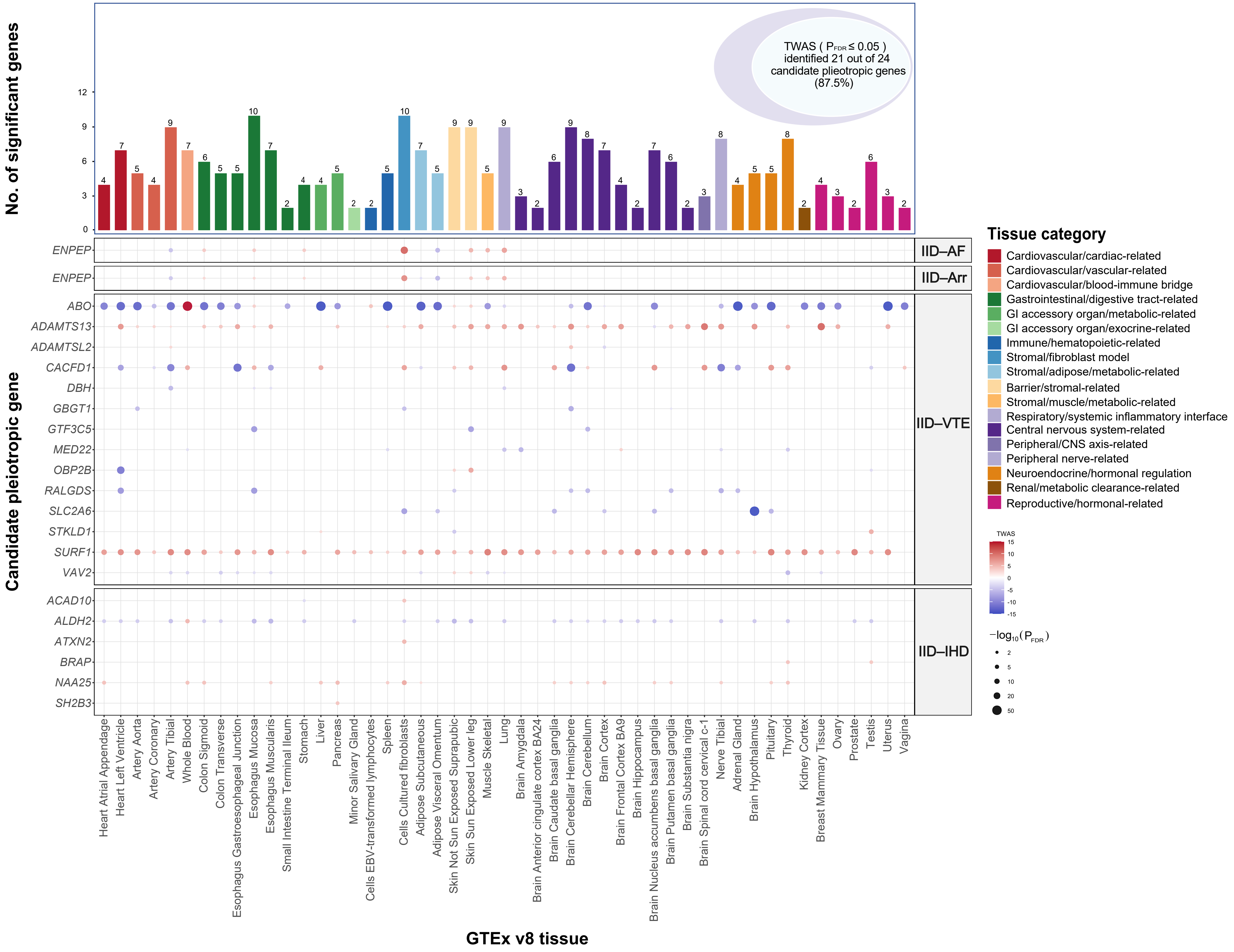

No. of significant genes
TWAS ( $P_{FDR}$ ≤ 0.05 ) identified 21 out of 24 candidate pleiotropic genes (87.5%)
Candidate pleiotropic gene
ENPEP
ABO
ADAMTS13
ADAMTSL2
CACFD1
DBH
GBGT1
GTF3C5
MED22
OBP2B
RALGDS
SLC2A6
STKLD1
SURF1
VAV2
ACAD10
ALDH2
ATXN2
BRAP
NAA25
SH2B3
IID–AF
IID–Arr
IID–VTE
IID–IHD
GTEx v8 tissue
Heart Atrial Appendage
Heart Left Ventricle
Artery Aorta
Artery Coronary
Artery Tibial
Whole Blood
Colon Sigmoid
Colon Transverse
Esophagus Gastroesophageal Junction
Esophagus Mucosa
Esophagus Muscularis
Small Intestine Terminal Ileum
Stomach
Liver
Pancreas
Minor Salivary Gland
Cells EBV-transformed lymphocytes
Spleen
Cells Cultured fibroblasts
Adipose Subcutaneous
Adipose Visceral Omentum
Skin Not Sun Exposed Suprapubic
Skin Sun Exposed Lower leg
Muscle Skeletal
Lung
Brain Amygdala
Brain Anterior cingulate cortex BA24
Brain Caudate basal ganglia
Brain Cerebellar Hemisphere
Brain Cerebellum
Brain Cortex
Brain Frontal Cortex BA9
Brain Hippocampus
Brain Nucleus accumbens basal ganglia
Brain Putamen basal ganglia
Brain Substantia nigra
Brain Spinal cord cervical c-1
Nerve Tibial
Adrenal Gland
Brain Hypothalamus
Pituitary
Thyroid
Kidney Cortex
Breast Mammary Tissue
Ovary
Prostate
Testis
Uterus
Vagina
Tissue category
Cardiovascular/cardiac-related
Cardiovascular/vascular-related
Cardiovascular/blood-immune bridge
Gastrointestinal/digestive tract-related
GI accessory organ/metabolic-related
GI accessory organ/exocrine-related
Immune/hematopoietic-related
Stromal/fibroblast model
Stromal/adipose/metabolic-related
Barrier/stromal-related
Stromal/muscle/metabolic-related
Respiratory/systemic inflammatory interface
Central nervous system-related
Peripheral/CNS axis-related
Peripheral nerve-related
Neuroendocrine/hormonal regulation
Renal/metabolic clearance-related
Reproductive/hormonal-related
TWAS
$-\log_{10}(P_{FDR})$
