## Supplemental Fig S10, and will be used for the link to the file on the preprint site. for "Shared genetic architecture between digestive infectious diseases and cardiovascular diseases: a genome-wide cross-trait analysis"

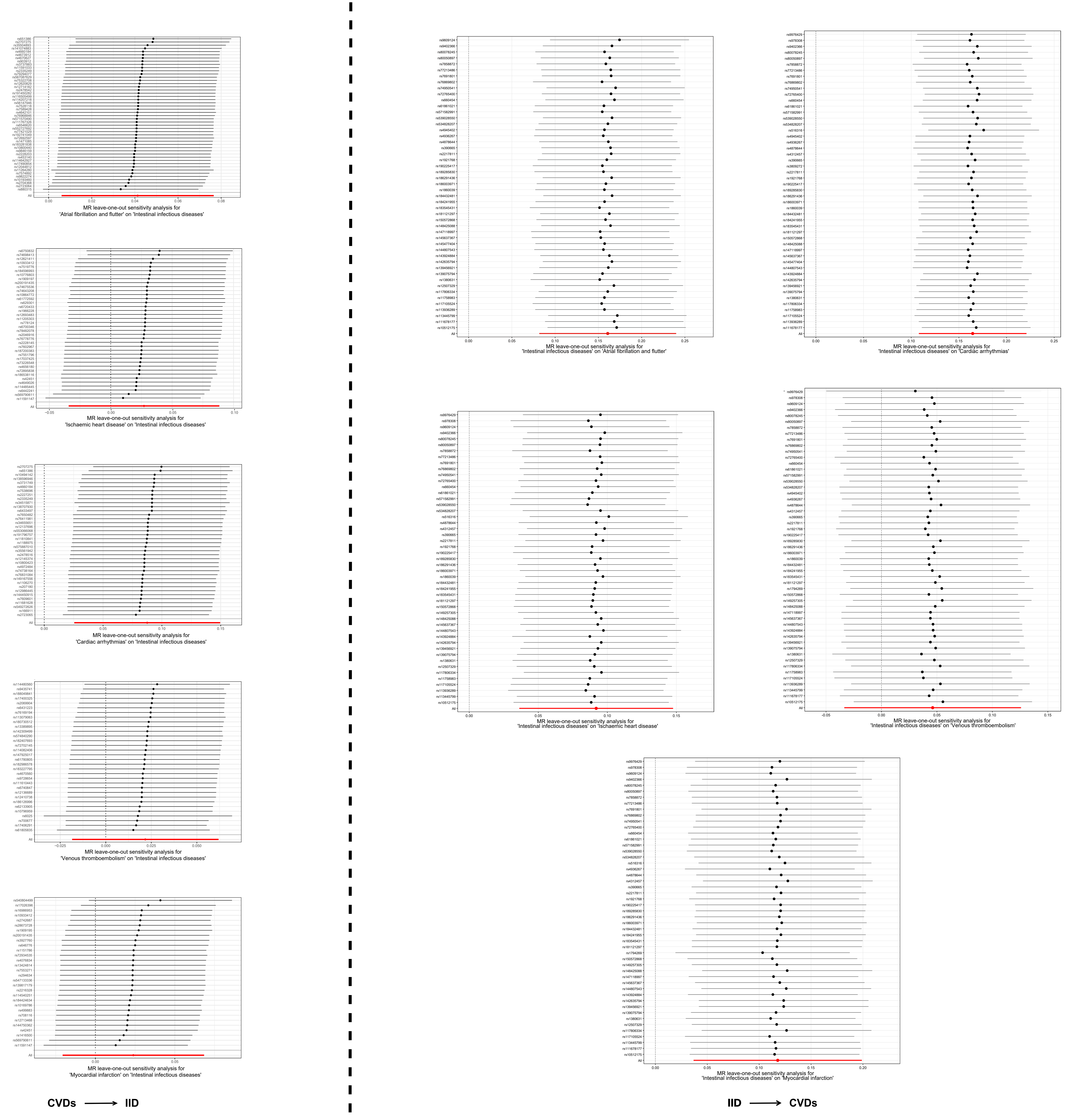
MR leave-one-out sensitivity analysis for
'Atrial fibrillation and flutter' on 'Intestinal infectious diseases'
MR leave-one-out sensitivity analysis for
'Ischaemic heart disease' on 'Intestinal infectious diseases'
MR leave-one-out sensitivity analysis for
'Cardiac arrhythmias' on 'Intestinal infectious diseases'
MR leave-one-out sensitivity analysis for
'Venous thromboembolism' on 'Intestinal infectious diseases'
MR leave-one-out sensitivity analysis for
'Myocardial infarction' on 'Intestinal infectious diseases'
MR leave-one-out sensitivity analysis for
'Intestinal infectious diseases' on 'Atrial fibrillation and flutter'
MR leave-one-out sensitivity analysis for
'Intestinal infectious diseases' on 'Cardiac arrhythmias'
MR leave-one-out sensitivity analysis for
'Intestinal infectious diseases' on 'Ischaemic heart disease'
MR leave-one-out sensitivity analysis for
'Intestinal infectious diseases' on 'Venous thromboembolism'
MR leave-one-out sensitivity analysis for
'Intestinal infectious diseases' on 'Myocardial infarction'
CVDs → IID
IID → CVDs
