## Supplemental Fig S11, and will be used for the link to the file on the preprint site. for "Shared genetic architecture between digestive infectious diseases and cardiovascular diseases: a genome-wide cross-trait analysis"

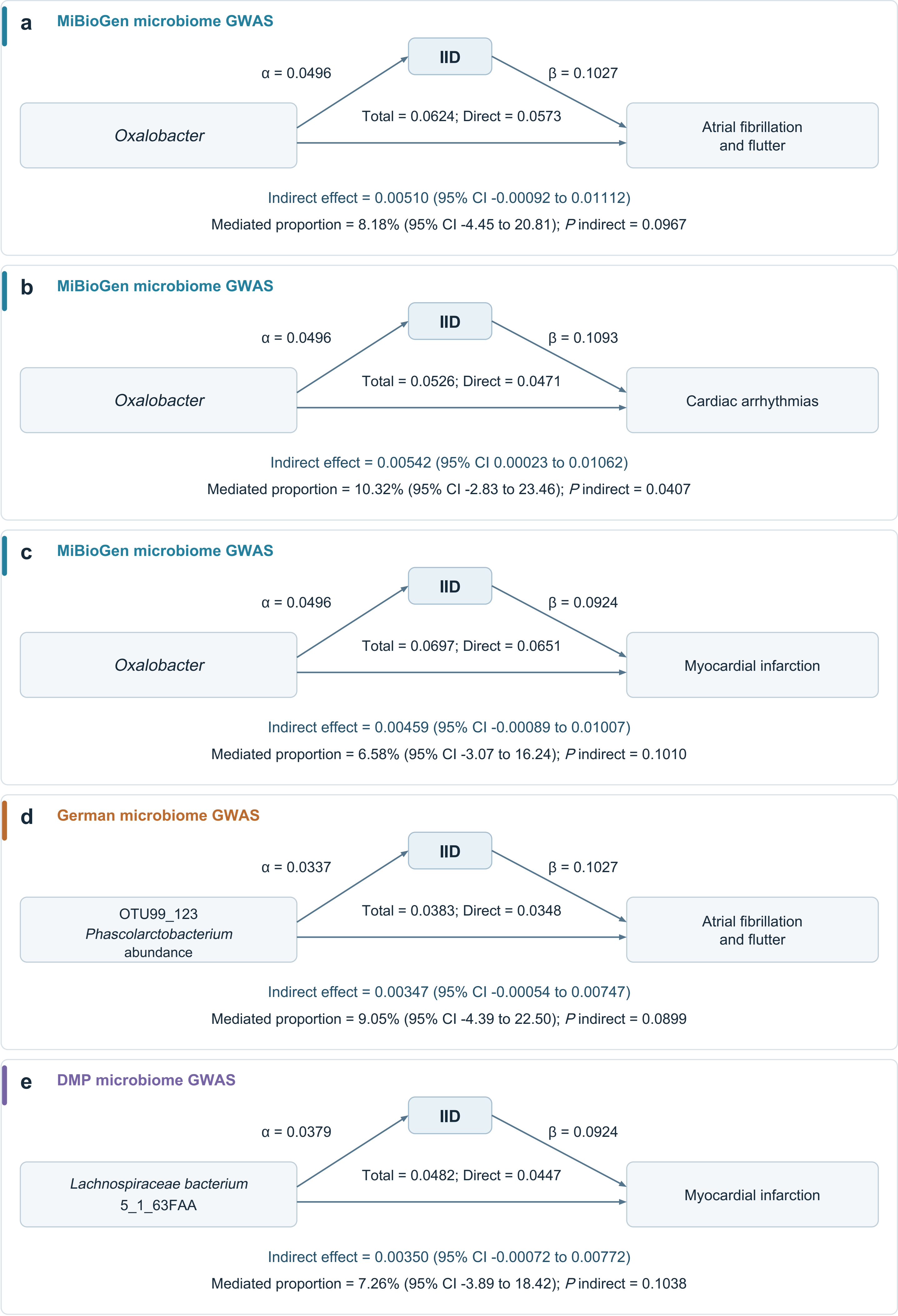
a
MiBioGen microbiome GWAS
IID
α = 0.0496
β = 0.1027
Oxalobacter
Total = 0.0624; Direct = 0.0573
Atrial fibrillation and flutter
Indirect effect = 0.00510 (95% CI -0.00092 to 0.01112)
Mediated proportion = 8.18% (95% CI -4.45 to 20.81); P indirect = 0.0967
b
MiBioGen microbiome GWAS
IID
α = 0.0496
β = 0.1093
Oxalobacter
Total = 0.0526; Direct = 0.0471
Cardiac arrhythmias
Indirect effect = 0.00542 (95% CI 0.00023 to 0.01062)
Mediated proportion = 10.32% (95% CI -2.83 to 23.46); P indirect = 0.0407
c
MiBioGen microbiome GWAS
IID
α = 0.0496
β = 0.0924
Oxalobacter
Total = 0.0697; Direct = 0.0651
Myocardial infarction
Indirect effect = 0.00459 (95% CI -0.00089 to 0.01007)
Mediated proportion = 6.58% (95% CI -3.07 to 16.24); P indirect = 0.1010
d
German microbiome GWAS
IID
α = 0.0337
β = 0.1027
OTU99_123 Phascolarctobacterium abundance
Total = 0.0383; Direct = 0.0348
Atrial fibrillation and flutter
Indirect effect = 0.00347 (95% CI -0.00054 to 0.00747)
Mediated proportion = 9.05% (95% CI -4.39 to 22.50); P indirect = 0.0899
e
DMP microbiome GWAS
IID
α = 0.0379
β = 0.0924
Lachnospiraceae bacterium 5_1_63FAA
Total = 0.0482; Direct = 0.0447
Myocardial infarction
Indirect effect = 0.00350 (95% CI -0.00072 to 0.00772)
Mediated proportion = 7.26% (95% CI -3.89 to 18.42); P indirect = 0.1038
